# Longitudinal study of immunity to SARS-CoV2 in Ocrelizumab-treated multiple sclerosis patients up to 2 years after COVID-19 vaccination

**DOI:** 10.1101/2024.01.23.24301671

**Authors:** Ilya Kister, Ryan Curtin, Amanda L. Piquet, Tyler Borko, Jinglan Pei, Barbara L Banbury, Tamar E. Bacon, Angie Kim, Michael Tuen, Yogambigai Velmurugu, Samantha Nyovanie, Sean Selva, Marie I. Samanovic, Mark J. Mulligan, Yury Patskovsky, Jessica Priest, Mark Cabatingan, Ryan C. Winger, Michelle Krogsgaard, Gregg J. Silverman

**Affiliations:** NYU Multiple Sclerosis Comprehensive Care Center, Department of Neurology, New York University Grossman School of Medicine, New York, NY; Laura and Isaac Perlmutter Cancer Center and Department of Pathology, NYU Grossman School of Medicine, New York, NY; Rocky Mountain MS Center, University of Colorado School of Medicine, Aurora, CO; Genentech, Inc., South San Francisco, CA; Adaptive Biotechnologies, Seattle, WA, USA; NYU Langone Vaccine Center and Department of Medicine, NYU Grossman School of Medicine, New York, NY; Division of Rheumatology, Department of Medicine, NYU Grossman School of Medicine, New York, NY

**Author notes:** Contributed equally (Michelle Krogsgaard, Gregg J. Silverman). **Corresponding authors:** Ilya Kister, Department of Neurology, New York University Grossman School of Medicine, 240 East 38^th^ St, Thirteenth Floor, New York, NY 10016, Michelle Krogsgaard, PhD, Department of Pathology, New York University Grossman School of Medicine, Smilow Research Center, 522 First Avenue, 701C, New York, NY 10016, Gregg Silverman, MD, Department of Medicine, NYU Grossman School of Medicine, 435 E 30th Street, Room 517, New York, NY 10016.

**Keywords:** multiple sclerosis, vaccinations, immunity

## Abstract

**Objectives:** 1 To plot the trajectory of humoral and cellular immune responses to the primary (two-dose) COVID-19 mRNA series and the third/booster dose in B-cell-depleted multiple sclerosis (MS) patients up to 2 years post-vaccination; 2. to identify predictors of immune responses to vaccination; and 3. to assess the impact of intercurrent COVID-19 infections on SARS CoV-2-specific immunity.

**Methods:** 60 Ocrelizumab-treated MS patients were enrolled from NYU (New York) and University of Colorado (Anschutz) MS Centers. Samples were collected pre-vaccination, and then 4, 12, 24, and 48 weeks post-primary series, and 4, 12, 24, and 48 weeks post-booster. Binding anti-Spike antibody responses were assessed with multiplex bead-based immunoassay (MBI) and electrochemiluminescence (Elecsys®, Roche Diagnostics), and neutralizing antibody responses with live-virus immunofluorescence-based microneutralization assay. Spike-specific cellular responses were assessed with IFNγ/IL-2 ELISpot (Invitrogen) and, in a subset, by sequencing complementary determining regions (CDR)-3 within T-cell receptors (Adaptive Biotechnologies). A linear mixed effect model was used to compare antibody and cytokine levels across time points. Multivariate analyses identified predictors of immune responses.

**Results:** The primary vaccination induced an 11-208-fold increase in binding and neutralizing antibody levels and a 3-4-fold increase in IFNγ/IL-2 responses, followed by a modest decline in antibody but not cytokine responses. Booster dose induced a further 3-5-fold increase in binding antibodies and 4-5-fold increase in IFNγ/IL-2, which were maintained for up to 1 year. Infections had a variable impact on immunity.

**Interpretation:** Humoral and cellular benefits of COVID-19 vaccination in B-cell-depleted MS patients were sustained for up to 2 years when booster doses were administered.

## INTRODUCTION

SARS CoV-2-specific immunity, which develops as a result of viral exposures and vaccinations, lowers the risk and severity of subsequent COVID-19 infections.^1,2^ However, immunocompromised individuals who have only a partial immune response to exogenous antigens are not fully immunoprotected after the relevant antigenic exposures.^3^ Thus, multiple sclerosis (MS) patients whose humoral immunity has been depressed with therapeutic B-cell depletion have a higher incidence of COVID-19 infections^4–7^ and COVID-19-related hospitalizations^8^ after they have been vaccinated compared to non-B-cell-depleted, vaccinated MS patients.^9^ At the same time, B-cell-depleted patients do benefit from vaccinations, as evidenced by their many-fold lower COVID-19 hospitalization rates following vaccination relative to the pre-vaccination epoch.^10,11^

To better understand the magnitude and durability of COVID-19 vaccine-induced immunity in patients treated with ocrelizumab, a B-cell depleting monoclonal therapy, we designed a prospective study—VIOLA (‘Vaccine-generated Immunity in Ocrelizumab-treated Patients: Longitudinal Assessments’, NCT04843774). The study assessed humoral and cellular immune responses to the primary (two-dose) COVID-19 mRNA vaccine series and to the third (booster) dose at multiple pre-specified time points up to 2 years from the initial vaccination.

The study period coincided with the emergence of the highly contagious Omicron variant, providing us with a unique opportunity to explore the impact of COVID-19 infections on SARS-CoV-2-specific immunity in vaccinated, ocrelizumab-treated patients.

## METHODS

### Inclusion and exclusion criteria

The study was approved by the Institutional Review Board of the NYU Grossman School of Medicine (New York). VIOLA was a prospective, two-center study of immunity against SARS-CoV-2 in MS patients receiving ocrelizumab (OCR) at the time of COVID-19 vaccination. All patients received neurologic care at the NYU MS Comprehensive Care Center in New York City (NYU) or the Rocky Mountain MS Center at the University of Colorado Anschutz Medical Center (UC-AMC). OCR infusions and COVID-19 vaccination were administered per standard of care.

The inclusion criteria were: clinician-diagnosed MS by the revised criteria;^12^ age 18 - 65 years; intent to undergo vaccination with mRNA COVID-19 vaccines (Pfizer-BioNTech/Comirnaty or Moderna/Spikevax) while on OCR; Expanded Disability Status Scale (EDSS) score ≤6.5; ability to provide written informed consent. Patients with prior COVID-19 were eligible. Baseline ‘SARS-CoV-2-infected’ status was determined based on a documented positive SARS CoV-2 polymerase chain reaction (PCR) or elevated anti-Spike antibody titer pre-vaccination, as previously described.^13^

The exclusion criteria were: prior vaccination for COVID-19; pregnancy, or planned pregnancy; breastfeeding; MS relapse within 3 months of study entry; known active infection; infection requiring hospitalization within 4 weeks of study entry; history of cancer, excluding localized skin cancers; immunodeficiency; concomitant disease requiring systemic corticosteroids or immunosuppressants; organ failure; active psychotic illness; active alcohol or drug abuse; immunosuppressives other than OCR; intravenous immunoglobulin or plasmapheresis within 3 months of study entry; prior alemtuzumab, cyclophosphamide, mitoxantrone, azathioprine, mycophenolate mofetil, cyclosporine, methotrexate, total body irradiation, or bone marrow transplantation; treatment with non-OCR anti-CD20 depleting agent within 6 months; severe hypogammaglobulinemia (IgG serum level was <300 mg/dL), lymphopenia (<750/mm^3^), or neutropenia (<1000/mm^3^).

### VIOLA visit schedule

All participants provided a blood sample within 26 weeks of the first vaccine dose and then 4 (+/- 7 days), 12 (+/- 14 days), 24 and 48 (+/- 28 days) weeks following the second vaccine dose. After VIOLA began, the CDC issued an advisory encouraging ‘booster’ (third dose) vaccination for immunocompromised individuals,^14^ and this recommendation was relayed to all VIOLA participants. Participants who opted for the booster dose (the ‘booster arm’) were invited to submit blood samples 4, 12, 24 and 48 weeks post-booster (with the same allowed time windows for sampling as above). Thus, participants were followed for up to 1 year after primary vaccination if they opted for no booster and up to 2 years if they opted for a booster.

At each study visit, a trained research coordinator interviewed patients using a structured instrument that elicited COVID-19 symptom history (per CDC clinical case definition) and information on any SARS-CoV-2 PCR and Antibody results/dates; COVID-19 treatments such as poly/monoclonal therapies; COVID-19-related and unrelated hospitalizations; MS relapses; adverse events related to OCR; new medications, procedures, and medical diagnoses. Electronic medical records were reviewed at each visit for relevant medical information. Study data were collected and managed using REDCap electronic data capture tool (https://www.project-redcap.org/) hosted at NYU Langone.

### Assessment of antibody responses to SARS CoV-2 vaccination and infection

Patients’ serologic responses to Spike protein were assessed by three different assays as previously described.^13^ Patients were tested at each time point for binding anti-Spike (Wuhan-strain) antibody levels using an Electrochemiluminescence immunoassay (Elecsys® platform, Roche Diagnostics GmbH, Mannheim, Germany) and a validated NYU proprietary custom Multi-epitope Bead-based Immunoassay (MBI).^15^ Live virus neutralization antibody activity (nAbs) against both Wuhan and Omicron SARS-CoV-2 strains were assessed using an immunofluorescence-based assay^16^ at baseline and 4-week post-primary vaccine and post-booster samples in all patients, and a subset of samples for the later time points. Samples collected after mono-/polyclonal antibody therapy for COVID-19 were excluded from antibody analyses.

Although VIOLA did not enroll healthy controls, samples collected 4 weeks after primary vaccination from 9 healthy, uninfected individuals were procured from the NYU Vaccine Center. The antibody levels in these subjects were measured using MBI and Wuhan/Omicron nAbs assays for reference.

### Assessment of cellular responses to COVID-19 vaccination and infection

T-cell responses to SARS-CoV-2 Spike protein were assessed in all patients and all time points with interferon-gamma (IFNγ) and IL-2 ELISpot (Invitrogen).^13^ IFNγ and IL-2 ELISpot values were calculated as spot-forming units (SFU) per 1 million cells. Additionally, for a subset of patients with available samples before and after booster or Omicron infection, antigen-specific T-cell immunity was assessed using Adaptive Immunosequencing T-MAP COVID (Adaptive Biotechnologies, Seattle, WA). This technology involves sequencing of the complementary determining regions (CDR)−3 within T-cell receptors (TCRs). T-cell responses specific to SARS CoV-2 were quantitated with COVID v3 classifier score.^17,18^

### Statistical analyses

Descriptive summaries of the results from the immunoassays were reported for continuous and categorical variables. Results that have heavily skewed distributions were normalized by log transformation. For continuous variables, mean, standard deviation (SD), median, and range were reported. For categorical variables, counts and percentages of patients with positive results were summarized. Correlation analyses were performed using Spearman correlation. Since the number of samples from each participant varied, a linear mixed effect model was used to compare antibody and cytokine levels between time points, where the comparison of 1^st^ and 3^rd^ time points was adjusted for time from vaccination to blood collection. We also carried out multivariate analyses to identify predictors of antibody and cellular responses 4 weeks after the primary vaccine series and 4 weeks after booster. The predictor variables are listed in the Results section.

For samples with immunosequencing data, peripheral T-cell repertoires were classified as ‘positive’ or ‘negative’ for detection of covid-specific T-cells using Adaptive’s COVID v3 classifier as described.^17,18^ In addition, the classifier reports a quantitative score that measures the magnitude of T-cell responses as a distance from pre-pandemic control populations. All statistical analyses of COVID v3 scores used nonparametric tests, including paired Wilcoxon signed rank and Spearman correlations.

## RESULTS

### Demographic and clinical characteristics of the VIOLA cohort

The cohort comprised 60 MS patients, 39 from NYU and 21 from the University of Colorado Anschutz Medical Center (CU-AMC). On average, each patient contributed samples at 6 time points (range: 2 – 8 samples per patient). The flowchart of VIOLA in **Figure 1** specifies the number of participants contributing blood samples at each time point. The enrollment period extended from January 2021 to November 2021. The last sample was collected in April 2023.

**Figure 1.**
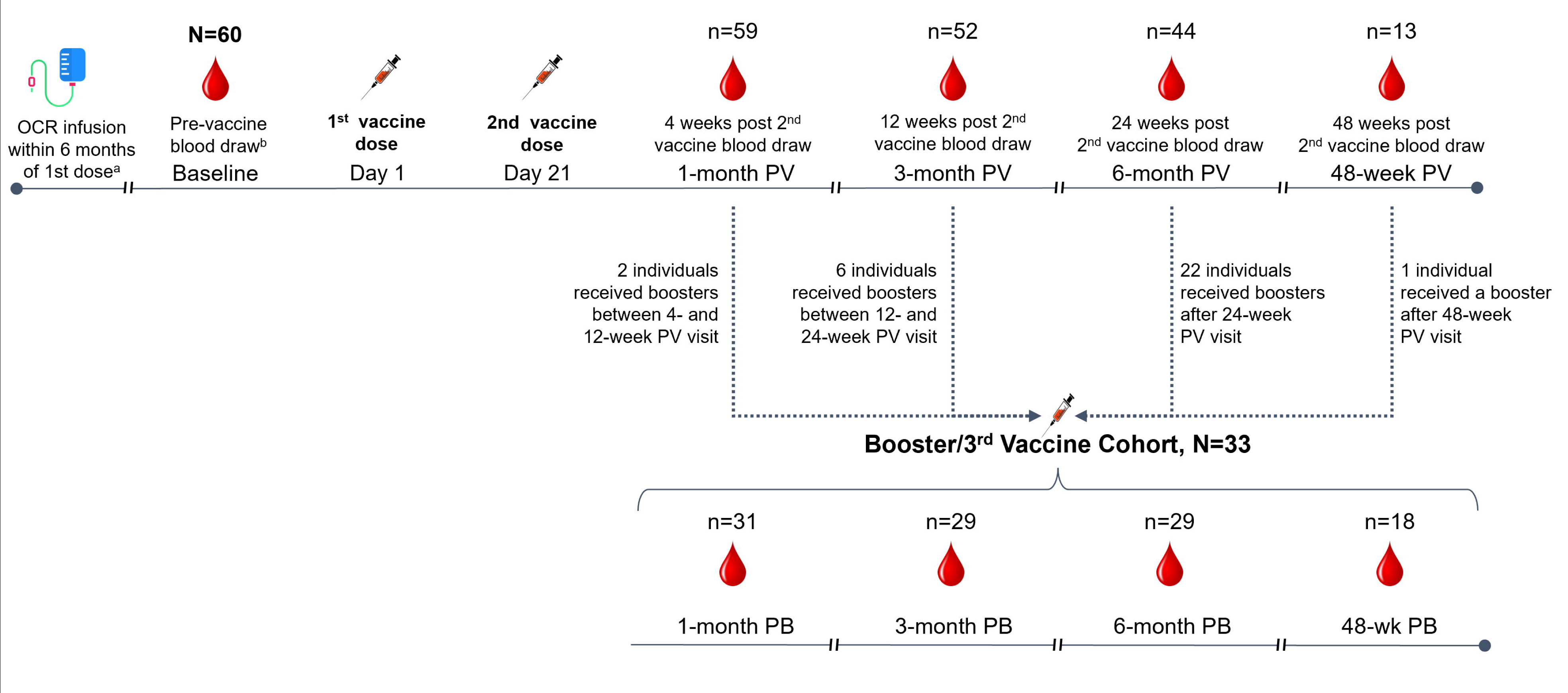
Study design. Bw, between; OCR, ocrelizumab; PB, post booster; pts; patients; PV, post vaccine; SOC, standard of care; wk, week. n=number of blood samples available for analyses at each timepoint. ^a^At least 2 weeks between last OCR and 1st vaccine. ^b^Vaccine anticipated within 4-6 weeks of baseline.

**Table 1** presents the demographic and clinical characteristics of the VIOLA cohort, as well as of NYU and CU-AMC subgroups. NYU and CU-AMC patients were similar with regard to all the demographic and clinical characteristics, except that more participants identified as non-White at NYU (76.9%) compared to CU-AMC (14.3%, p<0.001), a reflection of the different race/ethnic compositions of Centers’ catchment areas. 33 participants (55% of the VIOLA cohort) opted for booster during the study period. Demographic and clinical features of patients who received the booster were similar to those who did not (**Table S1**).

**Table 1.**
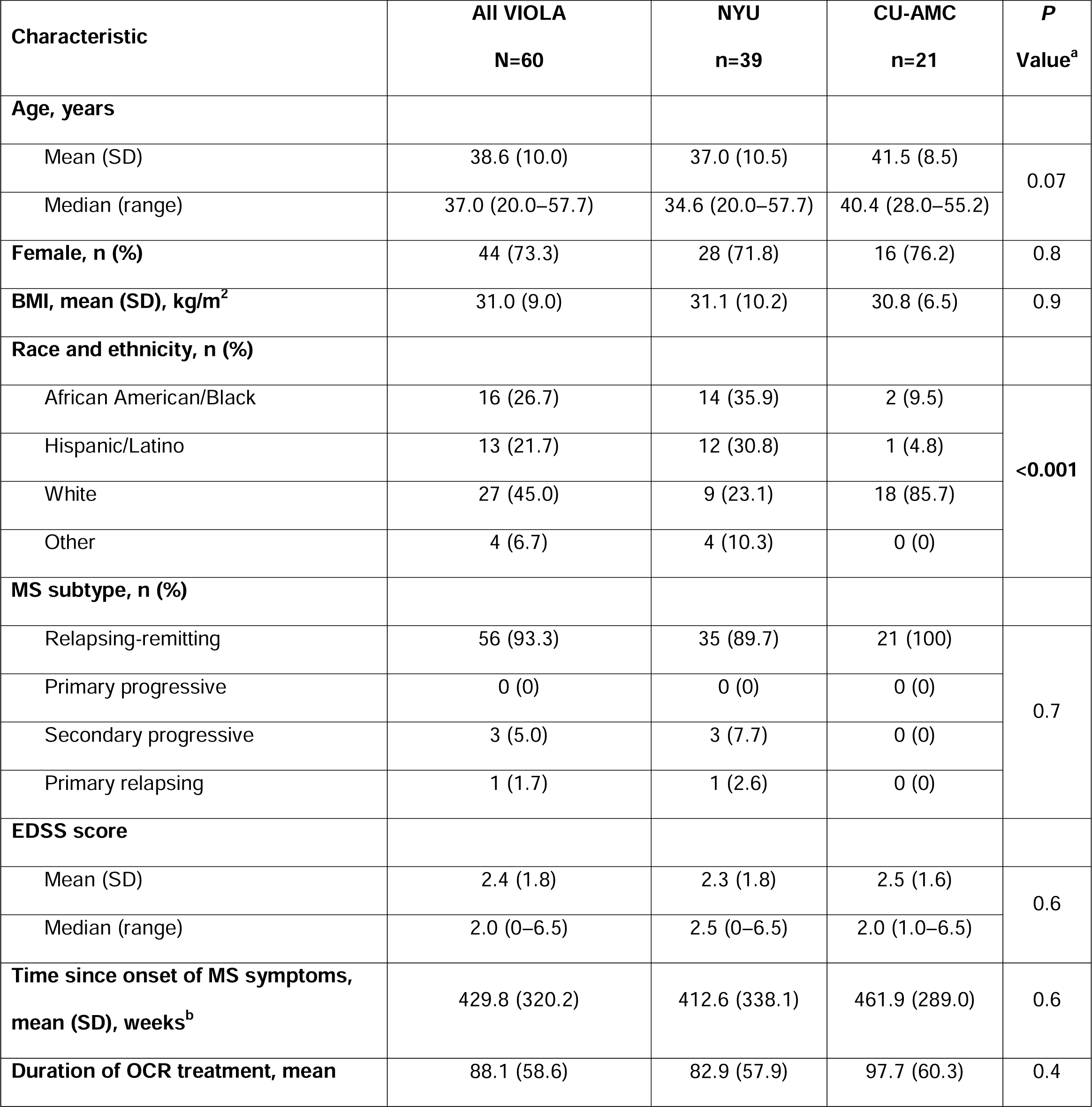
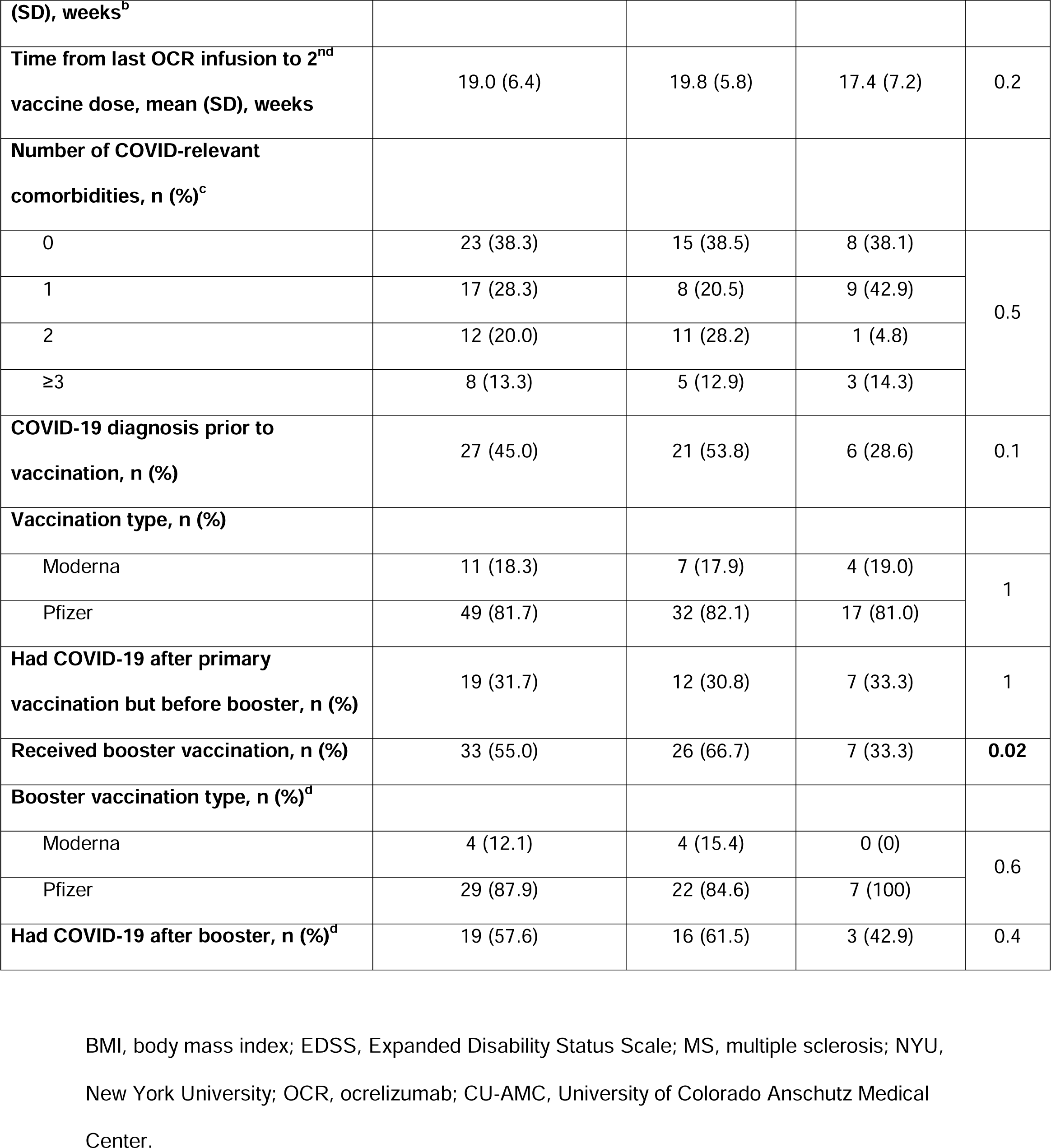
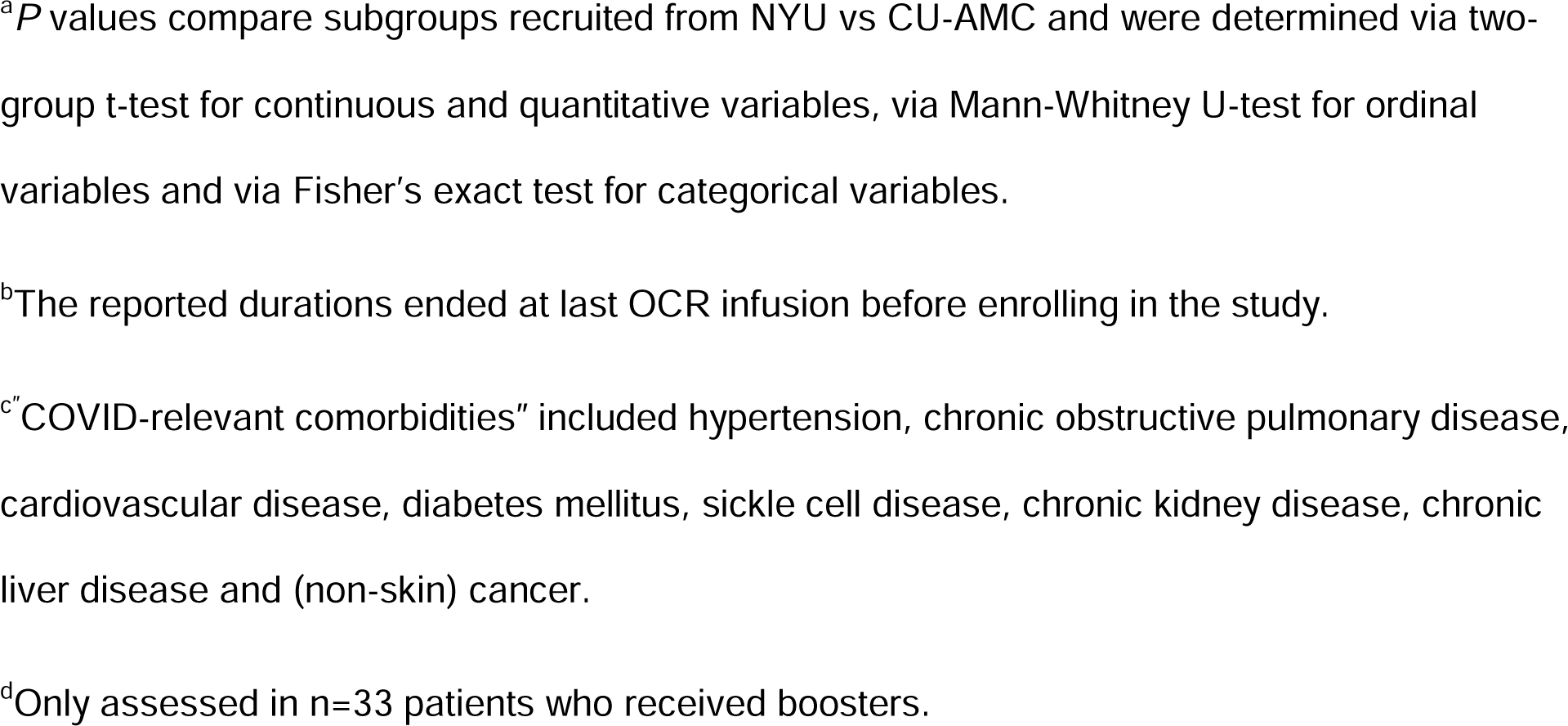
Demographics and clinical characteristics of the VIOLA cohort and of NYU and CU-AMC subsets.

| **Characteristic** | **All VIOLA**<br>**N=60** | **NYU**<br>**n=39** | **CU-AMC**<br>**n=21** | **P**<br>**Value<sup>a</sup>** |
| --- | --- | --- | --- | --- |
| **Age, years** |  |  |  |  |
| Mean (SD) | 38.6 (10.0) | 37.0 (10.5) | 41.5 (8.5) | 0.07 |
| Median (range) | 37.0 (20.0–57.7) | 34.6 (20.0–57.7) | 40.4 (28.0–55.2) |  |
| **Female, n (%)** | 44 (73.3) | 28 (71.8) | 16 (76.2) | 0.8 |
| **BMI, mean (SD), kg/m<sup>2</sup>** | 31.0 (9.0) | 31.1 (10.2) | 30.8 (6.5) | 0.9 |
| **Race and ethnicity, n (%)** |  |  |  |  |
| African American/Black | 16 (26.7) | 14 (35.9) | 2 (9.5) | <0.001 |
| Hispanic/Latino | 13 (21.7) | 12 (30.8) | 1 (4.8) |  |
| White | 27 (45.0) | 9 (23.1) | 18 (85.7) |  |
| Other | 4 (6.7) | 4 (10.3) | 0 (0) |  |
| **MS subtype, n (%)** |  |  |  |  |
| Relapsing-remitting | 56 (93.3) | 35 (89.7) | 21 (100) | 0.7 |
| Primary progressive | 0 (0) | 0 (0) | 0 (0) |  |
| Secondary progressive | 3 (5.0) | 3 (7.7) | 0 (0) |  |
| Primary relapsing | 1 (1.7) | 1 (2.6) | 0 (0) |  |
| **EDSS score** |  |  |  |  |
| Mean (SD) | 2.4 (1.8) | 2.3 (1.8) | 2.5 (1.6) | 0.6 |
| Median (range) | 2.0 (0–6.5) | 2.5 (0–6.5) | 2.0 (1.0–6.5) |  |
| **Time since onset of MS symptoms, mean (SD), weeks<sup>b</sup>** | 429.8 (320.2) | 412.6 (338.1) | 461.9 (289.0) | 0.6 |
| **Duration of OCR treatment, mean** | 88.1 (58.6) | 82.9 (57.9) | 97.7 (60.3) | 0.4 |
| (SD), weeks<sup>b</sup> |  |  |  |  |
| Time from last OCR infusion to 2<sup>nd</sup> vaccine dose, mean (SD), weeks | 19.0 (6.4) | 19.8 (5.8) | 17.4 (7.2) | 0.2 |
| Number of COVID-relevant comorbidities, n (%)<sup>c</sup> |  |  |  |  |
| 0 | 23 (38.3) | 15 (38.5) | 8 (38.1) | 0.5 |
| 1 | 17 (28.3) | 8 (20.5) | 9 (42.9) |  |
| 2 | 12 (20.0) | 11 (28.2) | 1 (4.8) |  |
| ≥3 | 8 (13.3) | 5 (12.9) | 3 (14.3) |  |
| COVID-19 diagnosis prior to vaccination, n (%) | 27 (45.0) | 21 (53.8) | 6 (28.6) | 0.1 |
| Vaccination type, n (%) |  |  |  |  |
| Moderna | 11 (18.3) | 7 (17.9) | 4 (19.0) | 1 |
| Pfizer | 49 (81.7) | 32 (82.1) | 17 (81.0) |  |
| Had COVID-19 after primary vaccination but before booster, n (%) | 19 (31.7) | 12 (30.8) | 7 (33.3) | 1 |
| Received booster vaccination, n (%) | 33 (55.0) | 26 (66.7) | 7 (33.3) | **0.02** |
| Booster vaccination type, n (%)<sup>d</sup> |  |  |  |  |
| Moderna | 4 (12.1) | 4 (15.4) | 0 (0) | 0.6 |
| Pfizer | 29 (87.9) | 22 (84.6) | 7 (100) |  |
| Had COVID-19 after booster, n (%)<sup>d</sup> | 19 (57.6) | 16 (61.5) | 3 (42.9) | 0.4 |

BMI, body mass index; EDSS, Expanded Disability Status Scale; MS, multiple sclerosis; NYU, New York University; OCR, ocrelizumab; CU-AMC, University of Colorado Anschutz Medical Center.
<sup>a</sup>*P* values compare subgroups recruited from NYU vs CU-AMC and were determined via two-group t-test for continuous and quantitative variables, via Mann-Whitney U-test for ordinal variables and via Fisher's exact test for categorical variables.
<sup>b</sup>The reported durations ended at last OCR infusion before enrolling in the study.
<sup>c</sup>"COVID-relevant comorbidities" included hypertension, chronic obstructive pulmonary disease, cardiovascular disease, diabetes mellitus, sickle cell disease, chronic kidney disease, chronic liver disease and (non-skin) cancer.
<sup>d</sup>Only assessed in n=33 patients who received boosters.

### COVID-19 infections among VIOLA participants

At the pre-vaccination baseline, 27 participants (45%) were classified as ‘previously SARS-CoV-2 infected’ based on positive PCR or elevated Spike antibodies (as defined in ^19^). Following vaccination, 36 patients (60%) experienced PCR-confirmed SARS-CoV-2 infections during the study period; two participants were infected twice with SARS-CoV-2. Half of the infections occurred after the second dose but before the booster (mean [SD] of 18.8 [11.9] weeks, range: 2.6 – 48.4 weeks, after the second dose), and half of the infections occurred after the booster (mean [SD] of 18.6 [12.7] weeks, range 1.0 - 47.3 weeks). Of the SARS-CoV-2 infections during the study period, 86% occurred within 3 months of the emergence of the Omicron variant (December 2021-March 2022). **Figure 2** provides timelines on OCR infusions, vaccinations, and infections for each participant, with the first dose of the vaccine as ‘time zero’.

**Figure 2.**
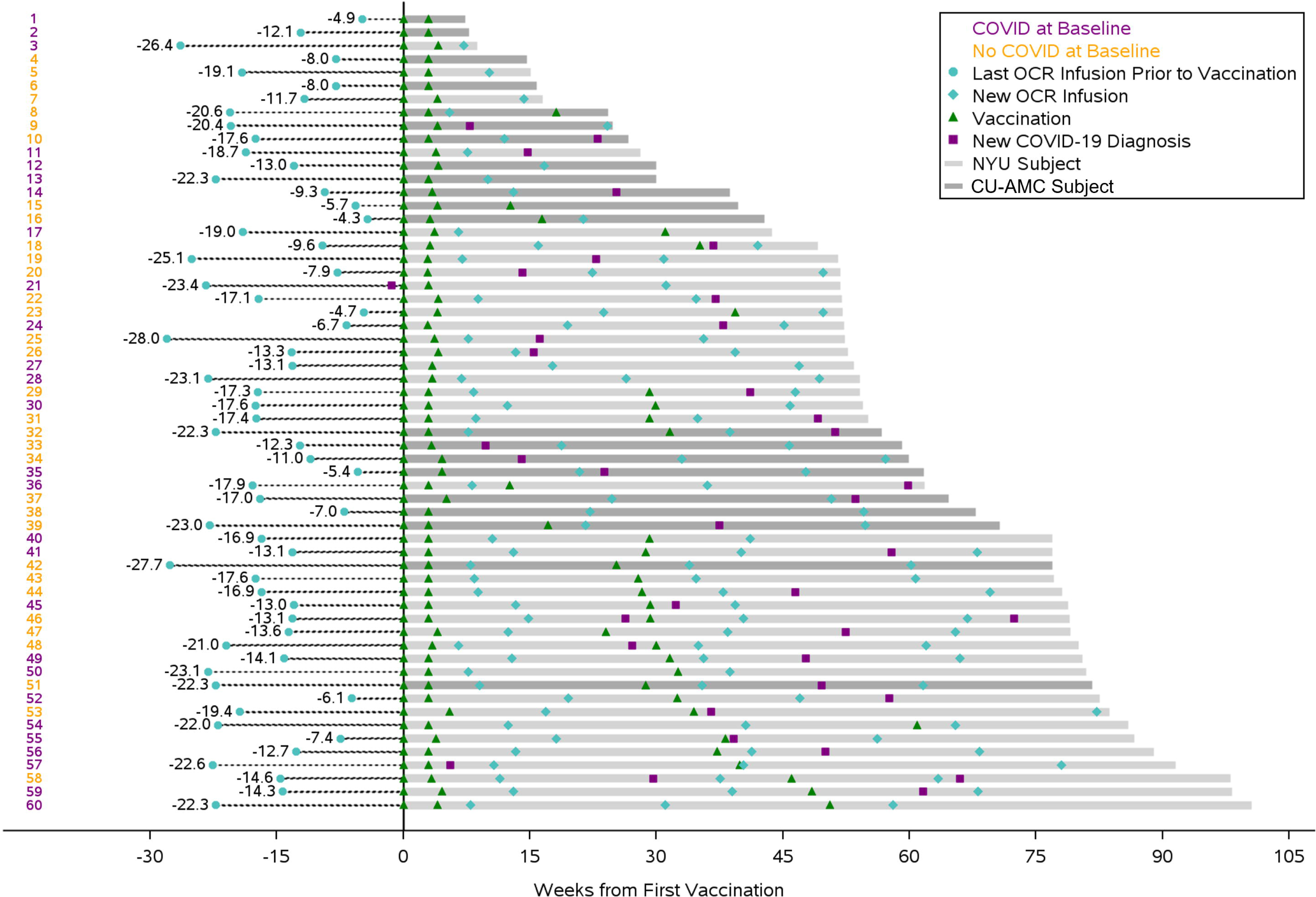
Timing of COVID-19 infections or vaccinations and ocrelizumab infusions in patients with MS. CU-AMC, University of Colorado Anschutz Medical Center; NYU, New York University; OCR, ocrelizumab. Study participants numbers shown in purple (left-most column) indicate that the respective participant was COVID-19-infected prior to vaccination, while those in orange were not infected prior to vaccination. Times from the last OCR infusion to vaccination is shown as dotted lines. ‘Time zero’ refers to the day of the first vaccine dose. Gray lines indicate the duration of participation. The darker gray shade refers to CU-AMC patients and the lighter gray shade refers to NYU patients.

Twelve patients infected during the study received mono/polyclonal anti-Spike antibody infusions/injections and were excluded from subsequent antibody analyses. Eight participants received COVID-19-specific antivirals. Three participants were hospitalized for COVID-19 during the study. One hospitalized patient underwent bronchoscopy, and another a lung biopsy, revealing SARS-CoV-2 virus in the lung parenchyma. No participant was intubated, admitted to the intensive care unit (ICU), suffered from stroke, heart attack, kidney failure, or died.

### The trajectory of anti-Spike binding antibody levels following primary vaccination

Primary vaccination induced an 11.1-fold increase in anti-Spike binding antibody levels as measured by MBI (p<0.001) (**Figure 3A1**) and a 207.5-fold increase by Elecsys (p<0.001) (**Figure 3B1**) at the 4-week time point. The antibody increases were significant in the subset of the previously SARS-CoV-2-infected as well as among the non-infected participants. The mixed effect model of antibody levels over 12 months following primary vaccination yielded a modest but significant decay (p=0.025 for MBI and p<0.001 for Elecsys). Despite the decay, the antibody levels remained significantly elevated relative to the pre-vaccine baseline for all time points up to 48 weeks (p<0.001 for all time points by MBI and p<0.05 for all time points by Elecsys).

**Figure 3.**
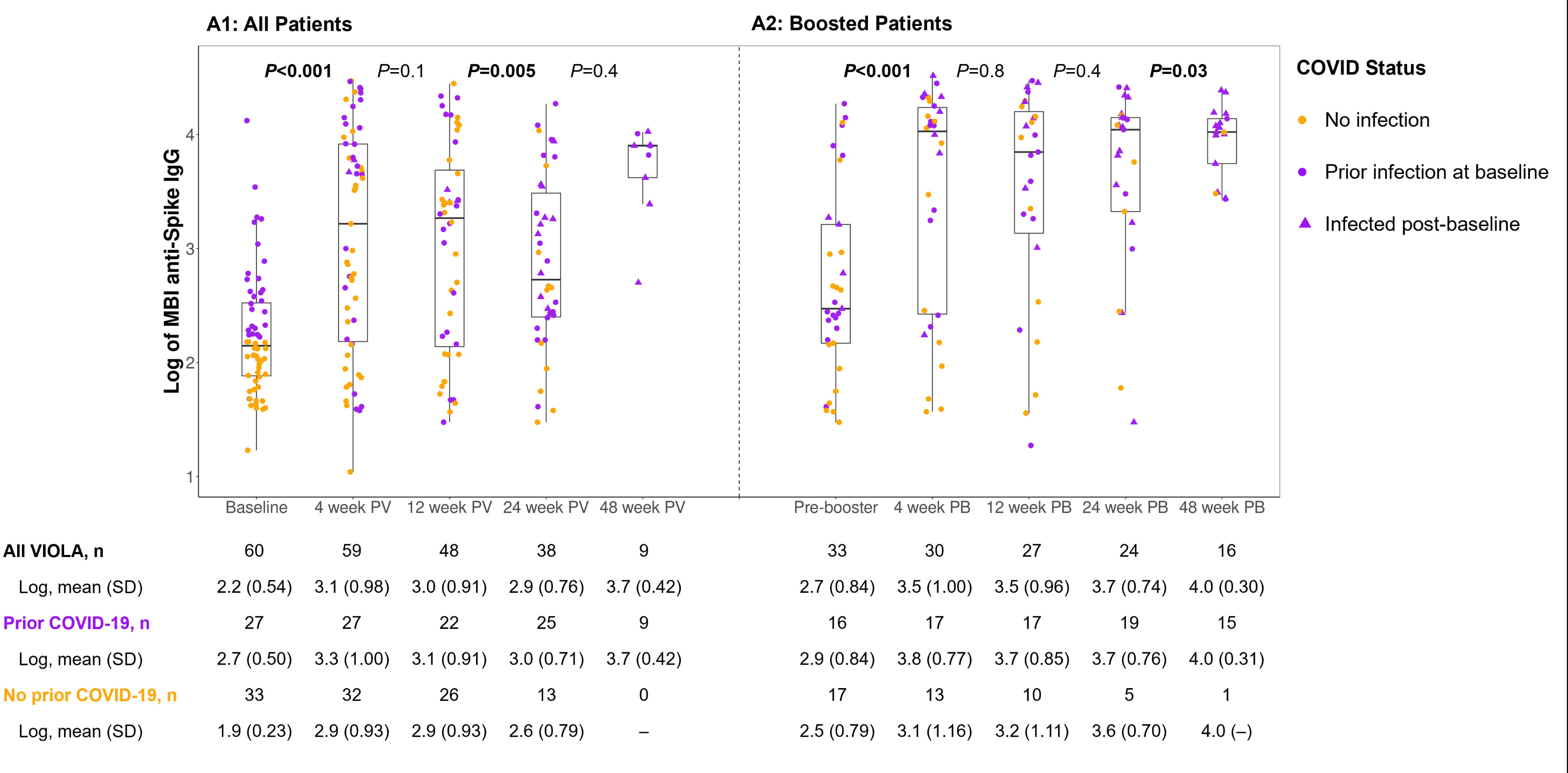
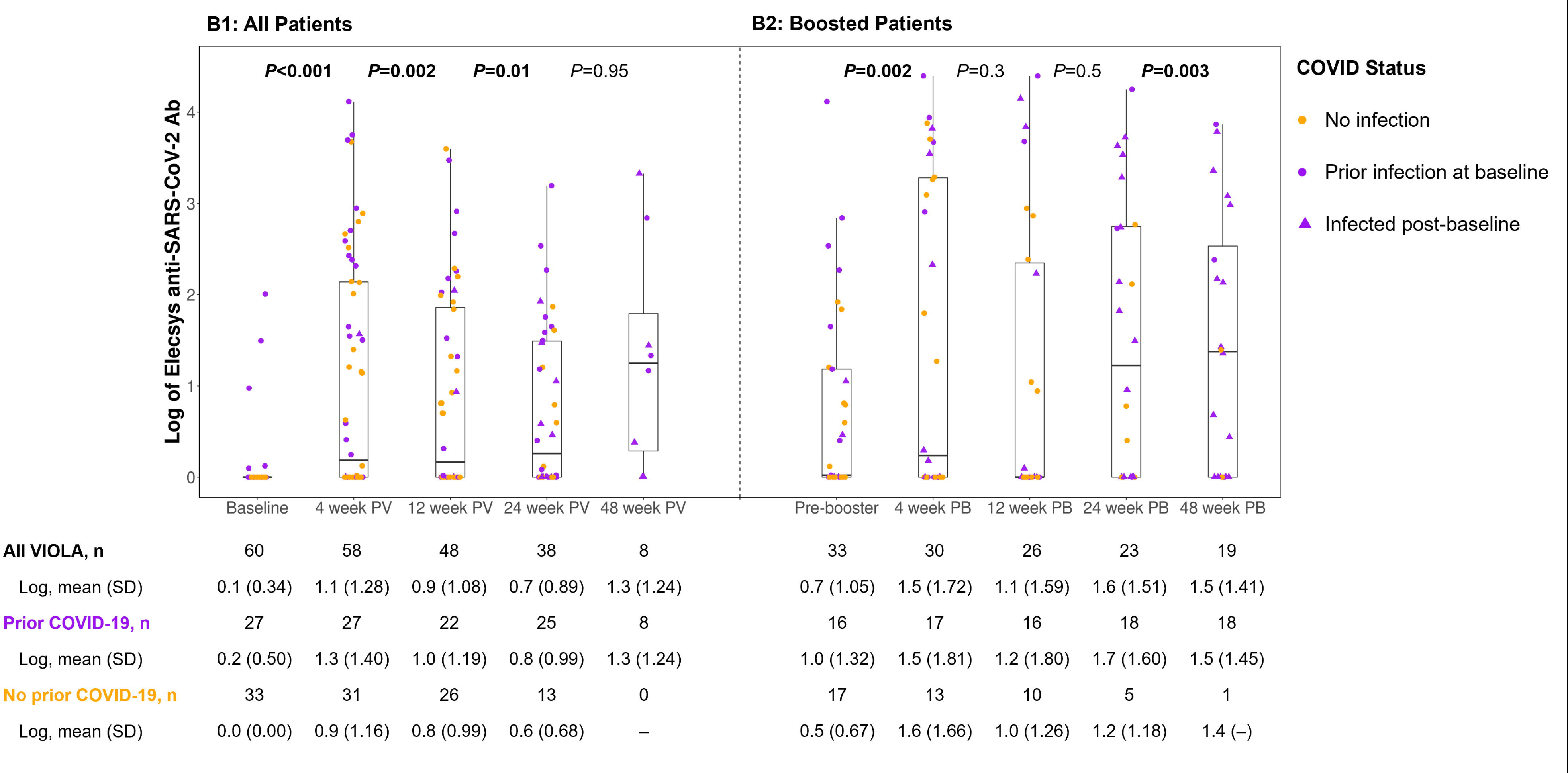
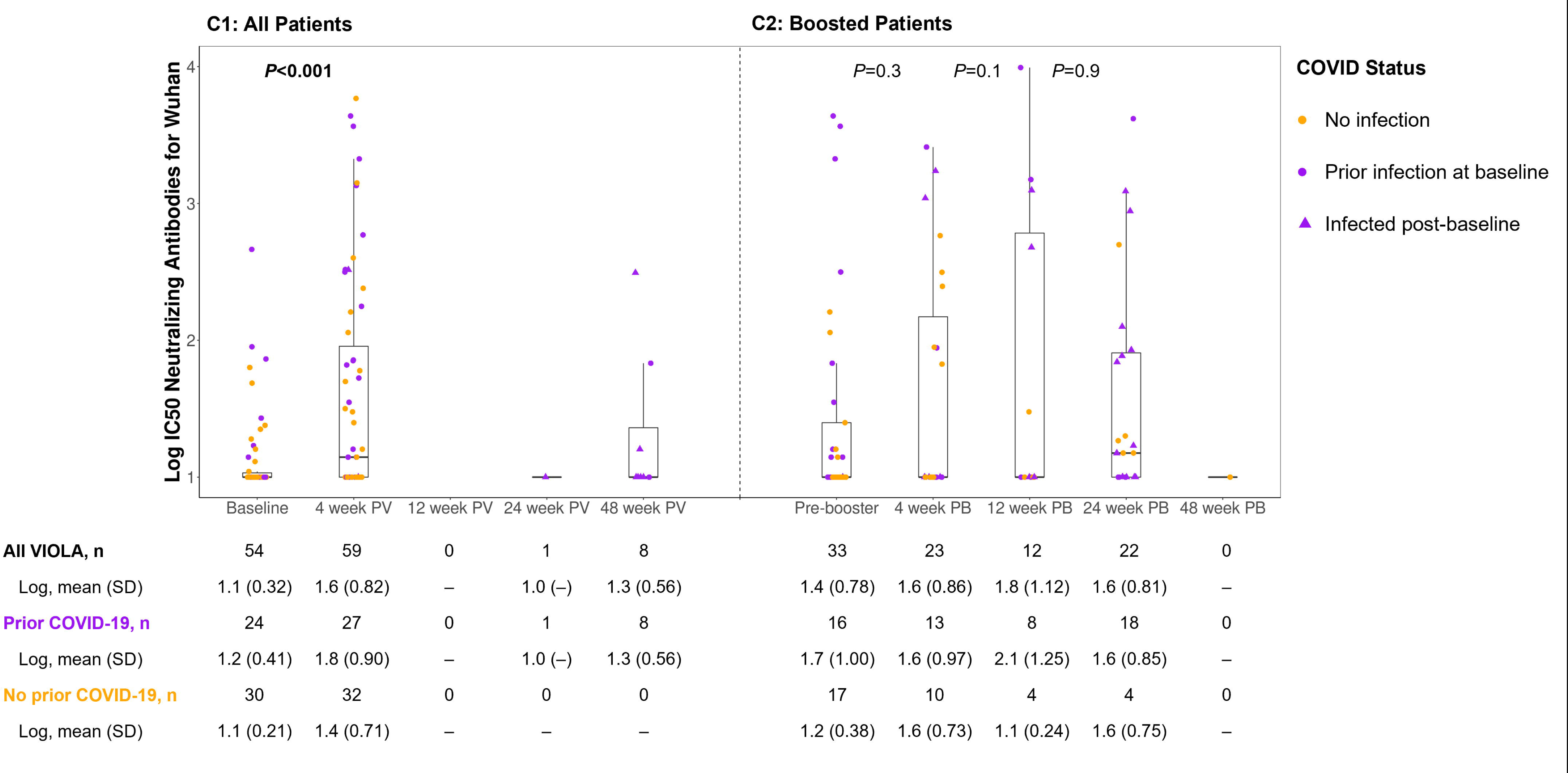
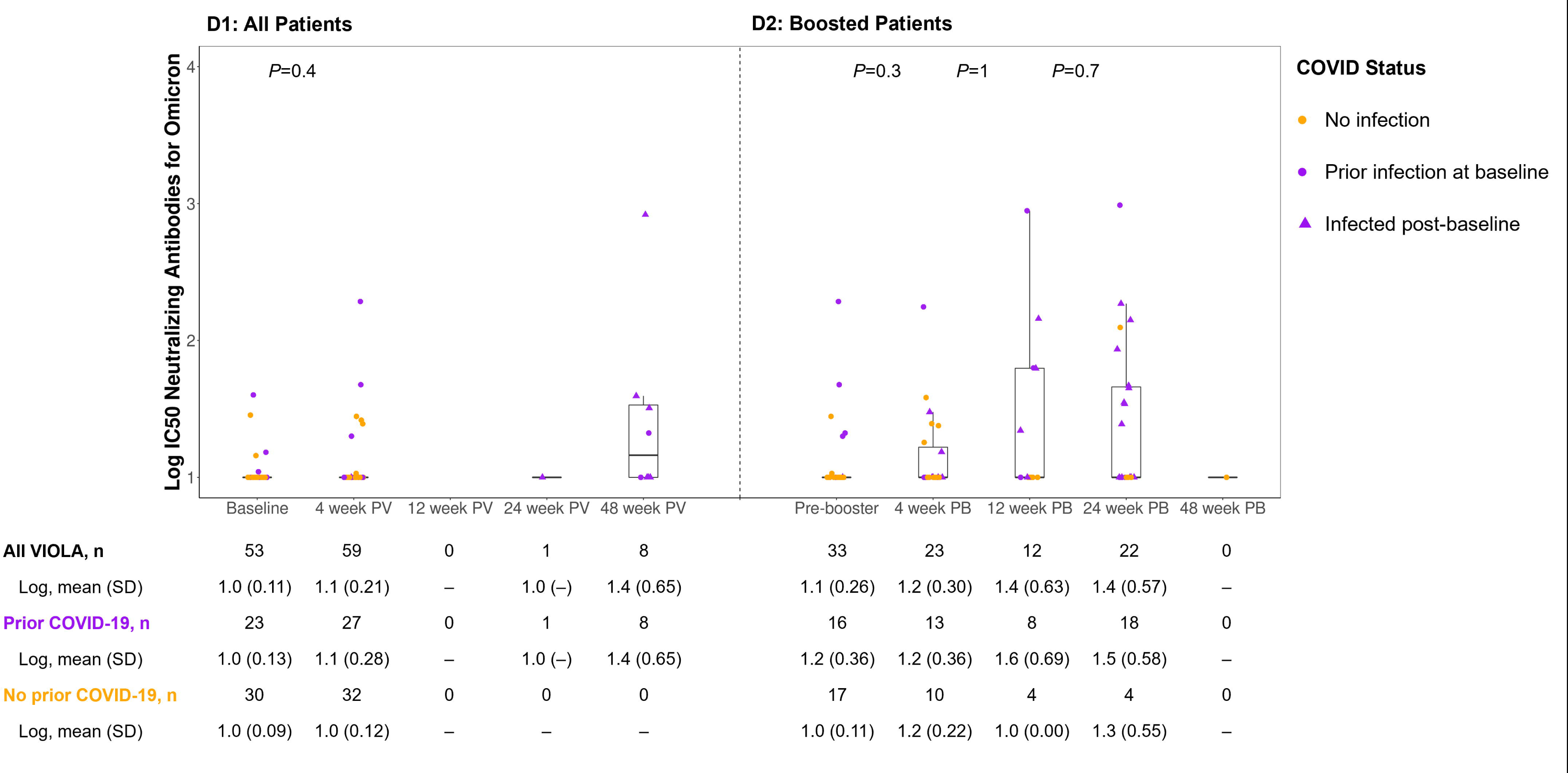
Post-vaccination (N=60) and post-booster (n=33) antibody responses as assessed by (A) MBI anti-Spike, (B) Elecsys anti-SARS-CoV-2, (C) Wuhan strain-neutralizing and (D) Omicron-strain neutralizing antibodies. PB, post booster; PV, post vaccine. Tables under each figure provide antibody levels for each time point for all patients, as well as for subsets of patients who were infected prior to the given time point (purple) and those who were not infected (orange). *P* values compare log-10 transformed values of neighboring time points, determined by paired t-tests.

For patients who experienced PCR-confirmed SARS-CoV-2 infection between the second dose and booster and did not receive poly/monoclonal therapy for COVID-19, we calculated the change in anti-Spike antibody levels in the last available pre-infection sample and the first available post-infection sample. No significant change in binding antibody levels was observed with either MBI (p=0.377) or Elecsys (p=0.846) in the 14 infected patients (mean time from infection to post-infection sample collection: 8.5 weeks, range 2 - 17 weeks).

### Anti-SARS CoV-2 neutralizing antibody responses following primary vaccination

Neutralizing antibodies (nAbs) to the Wuhan ancestral SARS-CoV-2 strain were tested at baseline and 4 weeks post-vaccination in all but one patient and a subset of patients at other time points (**Figure 3C1**). Primary COVID-19 vaccine series induced a 15.7-fold increase in Wuhan nAbs (p<0.001). The increase was observed among the previously infected subset as well as non-infected participants. Among the 8 patients for whom Wuhan nAbs were measured at week 48 post-vaccination as well as week 4, the antibody levels declined significantly between these two time points (p=0.029).

The highly infectious Omicron variant of SARS CoV-2 was first reported in the US on December 1, 2021, and, within weeks, it accounted for nearly all COVID-19 infections in the community (https://gisaid.org/hcov19-variants/). The first-generation mRNA vaccine was not designed for Omicron and offered reduced protection against this variant,^20,21^ which may explain why no post-vaccination increase in Omicron nAbs was observed in VIOLA (p=0.41; **Figure 3D1**). The correlation coefficients between Omicron nAbs and binding antibody levels were 0.31, p=0.013 for MBI and 0.47, p<0.001 for Omicron, which was numerically lower than between Wuhan nAbs and binding antibody levels (0.6, p<0.001 for MBI and 0.81 p<0.001 for Elecsys). Of the 8 patients with available samples at week 48, five of whom were infected with Omicron, nAbs to Omicron were 12-fold higher at week 48 relative to week 4, but this difference was not significant (p=0.114) (**Figure 3D1**).

### Predictors of antibody responses 4 weeks after primary vaccination

The multivariate model of antibody responses included sex, age, BMI, duration of OCR therapy, OCR infusion-to-vaccination time, pre-vaccine COVID-19 infection status, vaccine type, Center (NYU vs. CU-AMC) and race/ethnicity (white v. non-white). Non-White race/ethnicity predicted higher binding antibody responses on both assays (p<0.001 for MBI and Elecsys), while longer duration on OCR predicted lower antibody responses by MBI (p=0.007), Wuhan (p=0.026), and Omicron (p=0.036) nAbs, but not by Elecsys. Prior infection predicted less antibody increase by MBI (p=0.046) but not by the other assays.

Nearly all VIOLA participants were vaccinated within 6 months of infusion (mean [SD] of 15.6 [6.4] weeks, **Figure 2**), which may explain why the OCR infusion-to-vaccination interval did not correlate with the magnitude of antibody responses at week 4 post-vaccine. Vaccine type was not a predictor of antibody responses, but the statistical power of this analysis is limited due to the fact that only 15% of patients were vaccinated with Moderna/Spikevax.

### The trajectory of anti-Spike antibody levels following the booster dose

The mean period [SD] between the second and booster doses was 28 [10.3] weeks (**Figure 1**) among the 33 patients in the booster arm. Four weeks after the booster, a 3.2-fold increase (p<0.001) was detected by MBI (**Figure 3A2**) and a 5.1-fold increase (p=0.002) by Elecsys (**Figure 3B2**). The increases were significant in the subset of patients with prior infection. The mixed effect model showed no decay over the 48 weeks following booster on either MBI (p=0.23) or Elecsys (p=0.39) assays, and anti-Spike levels remained significantly higher at all post-booster time points relative to the pre-booster time point (p < 0.001 for MBI (**Figure 3A2**) and p<0.05 for Elecsys (**Figure 3B2**). The booster did not induce a significant increase in Wuhan (**Figure 3C2**) or Omicron NAbs (**Figure 3D2**).

Among the 15 patients who were infected following the booster and were not treated with COVID-19 poly/monoclonals, binding antibody and Wuhan neutralizing levels did not increase following the infection (MBI, p=0.201; Elecsys, p=0.838, Wuhan nAbs, p=0.5), while nAbs to Omicron increased significantly 2.3-fold (p=0.016). The mean period between pre- and post-infection samples among these 15 patients was 9.9 weeks (range 1.9 - 26.6 weeks).

### Predictors of 4-week antibody response to the booster dose

A multivariate model of antibody responses included the same set of predictor variables as for the post-primary vaccine model along with two additional variables: time from second to booster dose and antibody level change from baseline to 4-week post-primary vaccine. The significant predictors of stronger post-booster antibody responses were younger age (p=0.03) and shorter time from last OCR to vaccination (p=0.05) for MBI; shorter duration on OCR (p=0.01) for Elecsys; NYU study center (p=0.01), Pfizer vaccine type (p<0.001) and not having COVID before booster (p<0.001) for Wuhan NAbs; and lower BMI (p=0.002) and shorter time from the second vaccine to booster (p<0.001) for Omicron NAbs.

### Binding and neutralizing antibody responses to COVID-19 vaccine in healthy subjects from NYU Langone Vaccine Center

In 9 healthy subjects (age 38 ± 9.7 years, 55% female) with no history of COVID-19 and negative nucleocapsid antibody prior to vaccination, binding antibody titers measured by MBI at 4 weeks post-vaccine were 4.42 +/- 0.13 (log-transformed). For comparison, among the 32 uninfected VIOLA participants, levels by MBI at the same time point were 2.87 +/- 0.93. Only 3/32 (9%) VIOLA participants were within two standard deviations of the mean for the healthy subjects. These results are comparable with the French report in which 9% of B-cell repleted MS patients reached the minimum threshold of antibody considered as ‘adequate protection’ by local health authorities.^22^ Neutralizing antibody titers against Wuhan in healthy subjects were 3.13 +/- 0.41. Among uninfected VIOLA participants, Wuhan nAbs were 1.42 +/-0.71. Only 4/32 (12.5%) VIOLA participants were within two standard deviations of the mean for the healthy controls.

### The trajectory of Spike-specific cellular responses following the primary vaccination

Mean ELISPot-IFNγ values at week 4 post-vaccination increased 3.0-fold relative to baseline (p < 0.001) and remained unchanged at consecutive time points up to week 48 (**Figure 4A1**). A similar pattern was observed for ELISPot IL-2: a 3.7-fold increase from baseline to week 4 (p<0.001) and nonsignificant changes for the consecutive time points (**Figure 4B1**). The subset of patients with prior COVID-19 also had an increase in both IFNγ and IL-2 responses following vaccination. The mixed effect model showed no decay of IFNγ (p=0.86) or IL-2 (p=0.48) responses over 12 months, and both IFNγ (p<0.003 for all time points) and IL-2 (p<0.002 for all time points, except for week 48, p=0.079) remained significantly elevated relative to the pre-vaccine baseline. Our multivariate model did not identify any predictors of cellular immune response to vaccination.

**Figure 4.**
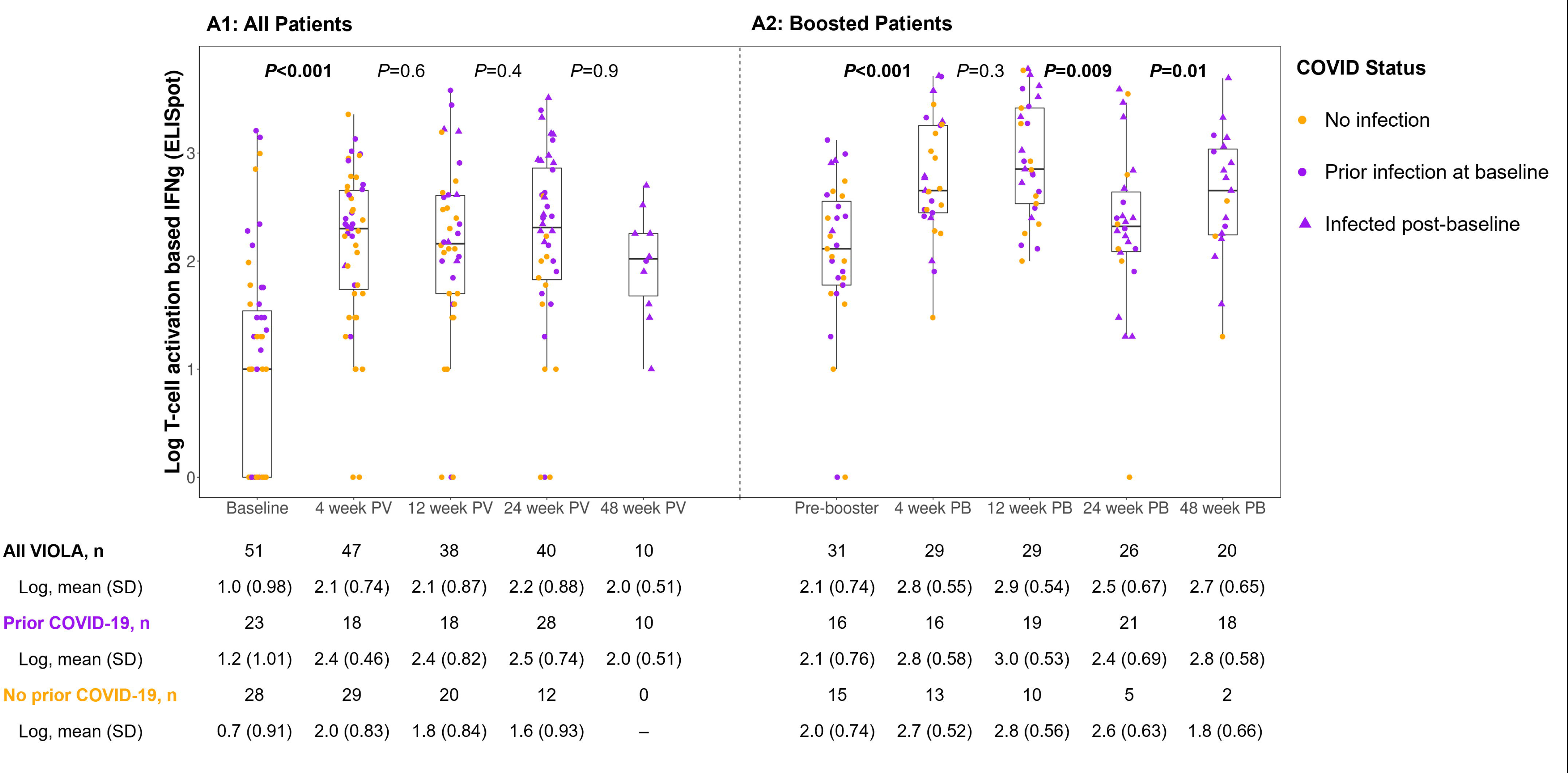
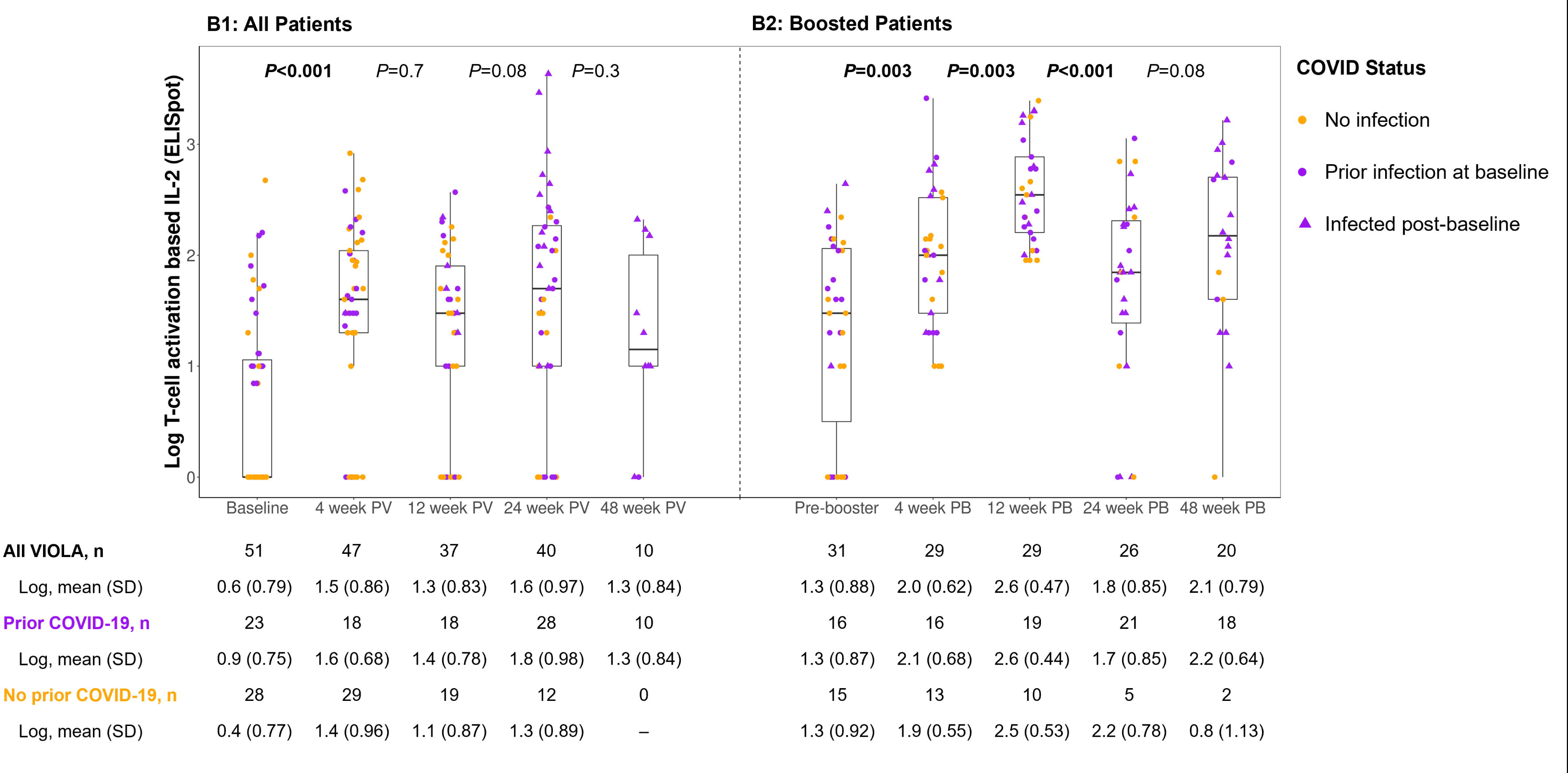
Post-vaccination (N=60) and post-booster (n=33) T-cell responses as assessed by (A) IFNγ and (B) IL-2. IFN, interferon; IL, interleukin; PB, post booster; PV, post vaccine. Tables under each figure provide cytokine levels for each time point for all patients, as well as for subsets of patients who were infected prior to the given time point (purple) and those who were not infected (orange). *P* values compare log-10 transformed values of neighboring time points, determined by paired t-tests.

To assess the impact of infection on cellular responses, we compared ELISpot IFNγ and IL-2 values pre- and post-infection in 14 patients who became infected between the second and booster vaccine doses. Among these patients, IFNγ values significantly increased 3.8-fold from pre- to post-infection time points (p=0.002), while IL-2 levels increased, non-significantly, 11.6-fold (p=0.127).

### The trajectory of Spike-specific cellular responses following booster

Booster induced a 4.6-fold increase (p<0.001) in IFNγ secreting cells on ELISpot assay (**Figure 4A2**) and a 3.7-fold increase (p=0.003) in IL-2-secreting cells (**Figure 4B2**) at week 4. The cellular responses remained significantly elevated above the pre-booster for all but one time point (p<0.001 for week 4, p<0.001 for week 12, p=0.027 for week 24 and p=0.003 for week 48 for IFNγ and p= 0.003 for week 4, p<0.001 for week 12, p=0.086 for week 24 and p=0.003 for week 48 for IL-2). In a multivariate model of cellular responses at 4 weeks post-booster that included the same predictor variables as for post-booster antibody responses, SARS-CoV-2-infected status before booster predicted lower IFNγ response (p=0.025). No predictors were identified for IL-2.

Among the 15 eligible patients who were SARS-CoV-2-infected following booster vaccination, ELISpot-IFNγ value decreased 2-fold from pre- to post-infection time points (p=0.053), while IL-2 decreased 3.3-fold (p=0.033).

### Immunosequencing of T-cell receptors before and after booster and Omicron infection

For TCR immunosequencing analyses, we selected 33 samples from 16 patients who have had either booster, Omicron or both. These samples yielded a total of 635,535 T-cells (range: 49,944-1,719,812); 426,955 unique T-cells (range: 40,489-857,569); with productive Simpson Clonality (a measure of the clone frequency distribution) of 0.031 (range: 0.003-0.139). These values were all within range of the general population (Adaptive; data on file). Nine patients contributed pre- and post-booster samples, 6 patients pre- and post-infection samples, and 1 patient – both. The timeline of vaccinations, infections, and OCR infusions relative to sample collection times for the 16 patients is shown in **Figure S1**.

The total number of T-cells, the number of productive T-cell rearrangements, and the COVID v3 classifier scores in vaccinated VIOLA participants who had Omicron and 20 vaccinated reference healthy controls who had Omicron from the Adaptive database did not differ with regard to any T-cell response characteristics, despite the younger age of the healthy subjects (**Figure 5**).

**Figure 5.**
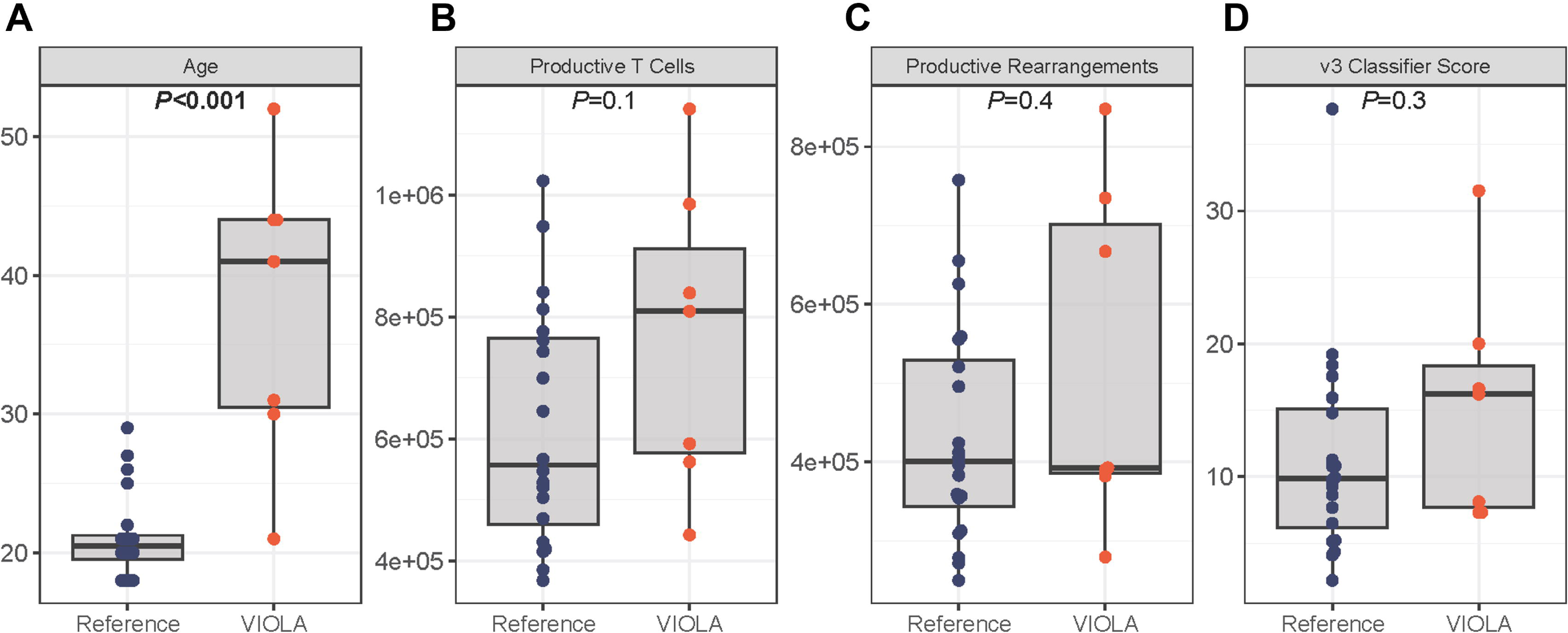
T cell characteristics of vaccinated VIOLA patients who had Omicron infection versus vaccinated reference healthy controls who had Omicron infection. *P* values were determined by Wilcoxon test.

COVID v3 scores of VIOLA participants did not correlate with their age, duration on OCR, time from OCR infusion to sample collection or time from vaccination, nor with levels of cellular (IFNγ and IL-2 ELISpot) or humoral (anti-Spike levels by MBI) immunity, with the exceptions of a trend for lower COVID v3 scores with longer time from vaccine in pre-booster/pre-Omicron samples and a trend for higher COVID v3 score and ELISpot-IFNγ levels in post-booster/post-Omicron samples (r=0.55, p = 0.023, **Figure 6**). COVID v3 scores in VIOLA participants before and after Omicron infection (n=7) and before and after booster (n=10) were unchanged (**Figure 7**).

**Figure 6.**
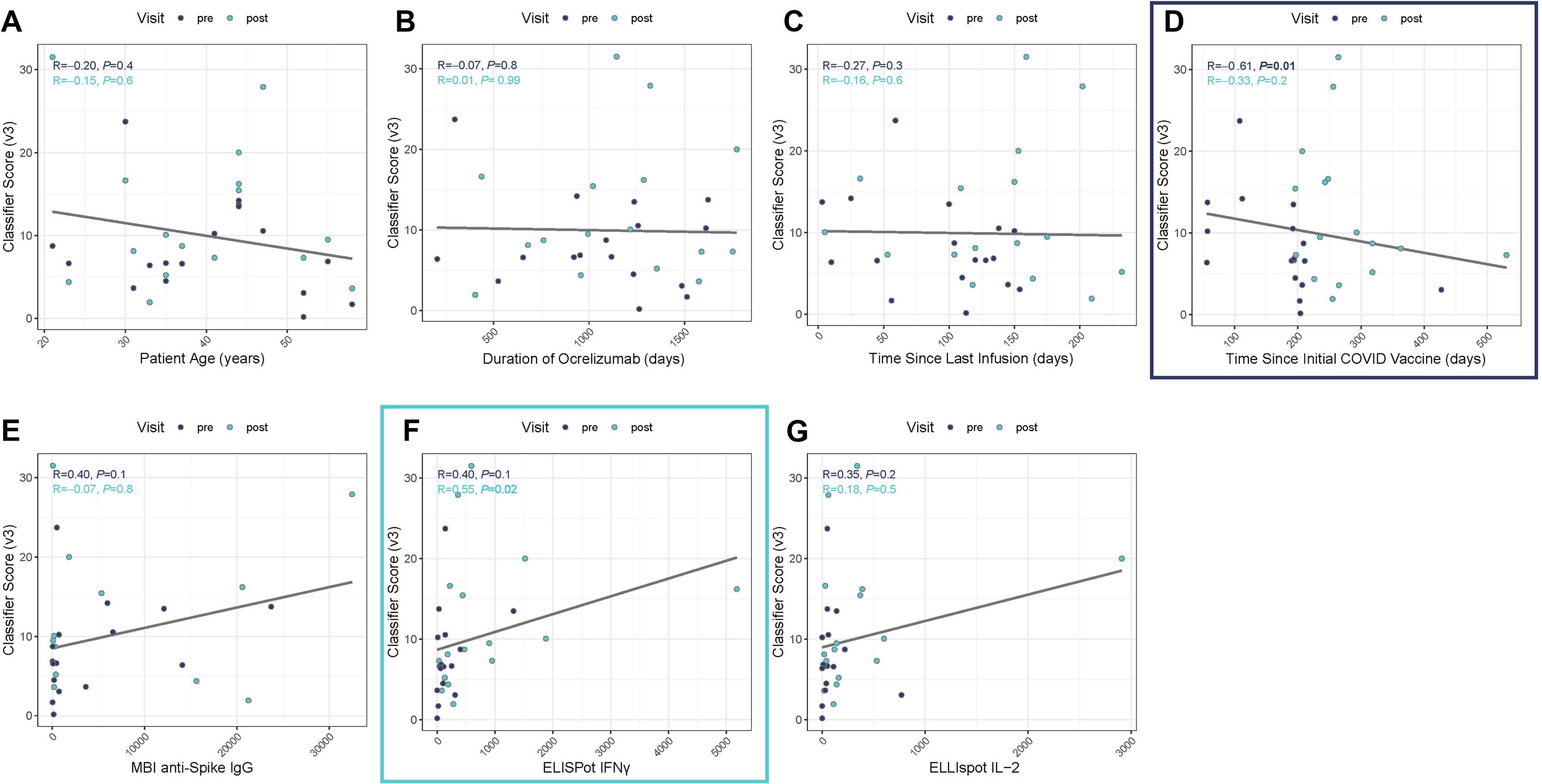
COVID V3 score correlations with participant characteristics and immune responses. IFN, interferon; Ig, immunoglobulin; IL, interleukin; MBI, multi-epitope bead-based immunoassay. ‘Pre’ corresponds to samples collected prior to booster or Omicron. ‘Post’ corresponds to samples collected after booster or Omicron. ‘Pre’ and ‘post’ samples are analyzed separately. R and *P* values were determined via Spearman correlation. Blue boxes indicate correlations that reached significance in either pre- or post-booster/Omicron subsets of samples.

**Figure 7.**
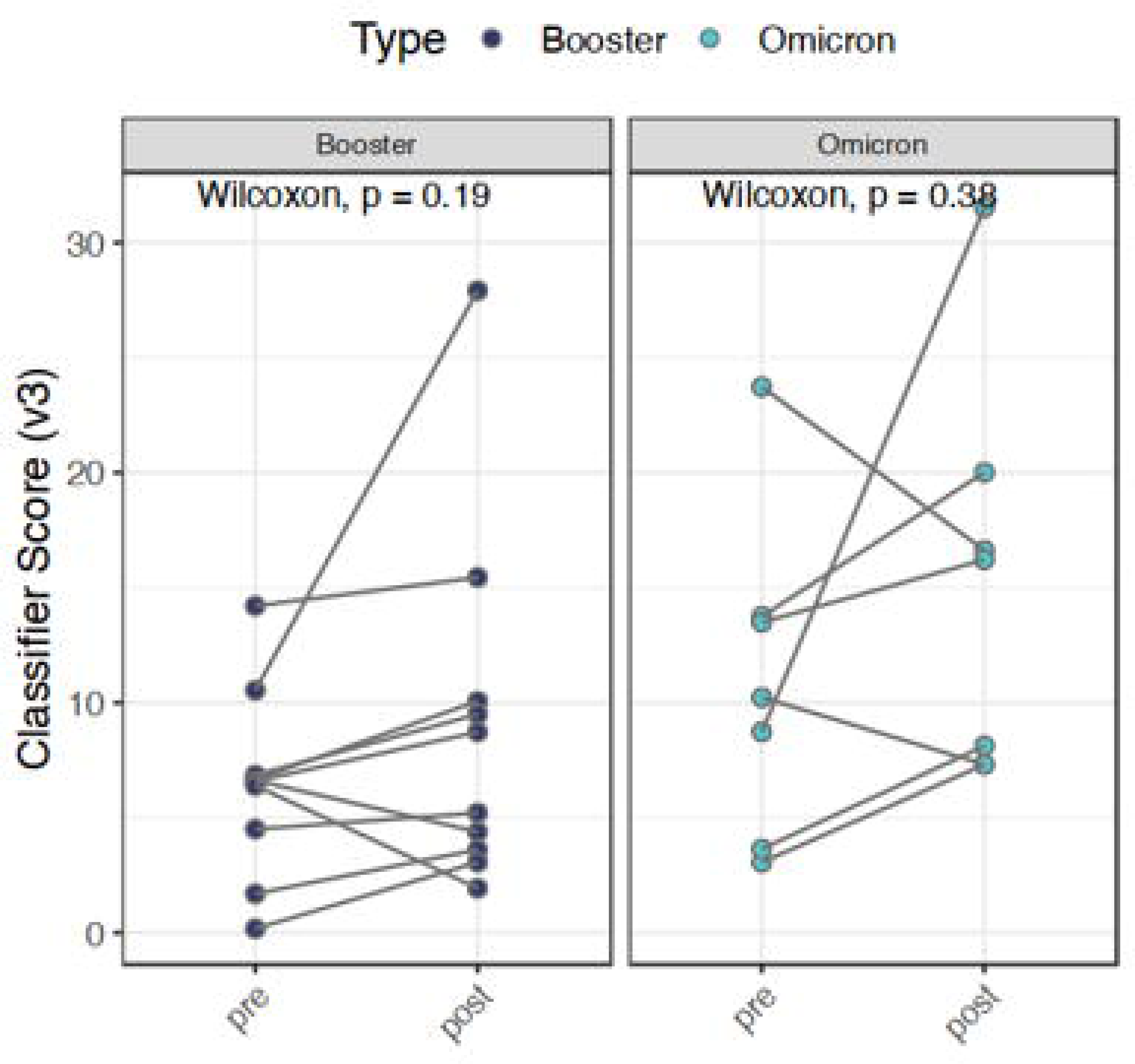
COVID V3 scores before and after^a^ booster or Omicron strain infection. *P* values were determined via Wilcoxon test. ^a^12 weeks after booster and at the next collection after COVID-19 infection.

## DISCUSSION

Primary mRNA COVID-19 vaccination series induced a highly significant increase in anti-Spike binding and Wuhan neutralizing antibody levels in both infected and uninfected VIOLA participants (Fig. 3A1, 3B1). Even so, the antibody levels in our patients were much lower than in the reference healthy subjects, and a minority of VIOLA participants did not develop any post-vaccine antibody responses, in line with prior studies.^11,19,22–27^ Binding and neutralizing antibody levels decayed over time but remained elevated above the pre-vaccine level up to 12 months following the primary vaccine. This is similar to what has been observed for the general population: anti-Spike antibody responses peak around 21–28 days after the second vaccine dose and then decline,^28^ presumably due to a decline in short-lived plasmablasts.^29^

The literature is conflicted with regard to the efficacy of boosters in B-cell depleted patients: some report an increase in antibody levels,^30,31^ and others do not.^32,33^ In VIOLA, the booster induced a significant 3-5-fold rise in binding antibodies but no significant rise in nAbs against the Wuhan strain. The lack of nAbs increase after booster is consistent with one prior study using live virus assay in which “neutralizing activity against tested SARS-CoV-2 variants remained low or undetectable” after third or fourth booster doses in aCD20-treated MS patients.^22^ An important novel finding of our study is that binding antibody levels remain stably elevated for up to 12 months after booster.

Neither primary vaccine nor booster raised Omicron nAbs levels, likely because the first-generation vaccines were designed against the Wuhan strain and not against Omicron and because ocrelizumab markedly attenuates humoral responses. In healthy subjects, primary vaccination with the first-generation vaccine was associated with a reduced nAb response to Omicron relative to Wuhan strain,^34^ while boosting with the newer-generation variant vaccine induces much higher nAbs against Omicron compared to booster with the prototype vaccine.^35^ The variant vaccines were FDA-approved in Sept 2023, and their impact on SARS-CoV-2 immunity in B-cell depleted population has not, to our knowledge, been reported.

Non-White race/ethnicity was a predictor of higher binding antibody levels following the primary vaccine series in all three assays. This finding is congruent with the emerging data on higher efficacy of the COVID-19 vaccines in persons of African and South Asian descent as compared to whites^36^ and with studies that document higher antibody responses to flu^37^ and rubella^38^ vaccines and faster B-cell repletion^39^ in persons of African descent. Patients who were treated with OCR for longer periods tended to have weaker antibody responses on all assays (except for Elecsys) in line with several prior studies.^22,40,41^ Longer infusion-to-vaccination time did not correlate with better antibody responses in VIOLA, likely because almost all VIOLA participants were vaccinated within 6 months of infusion (Table 1); in studies where the infusion-to-vaccination period was extended >6 months post-infusion, longer period consistently predicted better antibody response.^11,40^ Regarding post-booster antibody responses, the models yielded modest and variable predictors, with almost no overlap among the three antibody assays. We could not corroborate the results of a prior study^42^ that the magnitude of antibody response to the primary vaccine predicts the response to the booster.

Cellular responses to Spike protein following the primary series were significantly enhanced in both previously infected and uninfected subsets (Fig. 4A1, 4B1). Unlike antibody levels, cellular responses remained relatively stable up to one year following vaccination in VIOLA. This is consistent with studies in the healthy population, where cellular immunity to COVID-19 vaccination and infection tends to be more durable than antibody responses.^43^ Robust longitudinal cellular immunity in B-cell-depleted MS patients is a consistent finding across multiple studies.^26,33,44–47^ Booster further raised cytokine responses 4-5-fold. This was a similar-fold increase as after the primary vaccine. In contrast, antibody increase after booster was much less pronounced than after the primary series. The positive impact of booster on cellular immunity of B-cell depleted patients has been reported in most,^42,47,48^ but not all^33^ prior studies. An important finding of VIOLA is that cellular immune responses to Spike protein in VIOLA participants remained stably elevated up to one year after booster.

Immunosequencing of SARS CoV2-specific T-cell receptors (TCR)^17^ in the subset of 16 participants showed that B-cell depleted MS patients have comparable breadth and depth of Spike-specific T-cell clone as vaccinated healthy subjects from the reference dataset (Fig. 5), in agreement with Algu et al.^49^ COVID v3 score in VIOLA did not correlate with the duration of OCR time since last OCR infusion, or markers of functional cellular immunity, except for a positive correlation in post-booster/infection samples with IFNγ ELISpot. The overall lack of correlation may be due to the small sample size or to the fact that the two tests probe largely non-overlapping aspects of T-cell immunity (functional responses v. genetic rearrangements).

The highly contagious Omicron variant emerged several months after the start of VIOLA and infected most participants of the study, enabling us to investigate immune responses to infection in vaccinated, B-cell-depleted patients. Omicron-period infections did not raise binding or Wuhan neutralizing antibody levels but did raise Omicron-specific neutralizing antibody levels (Fig. 3D1). It is possible that binding antibody assays developed for the Wuhan Spike may not be effective for detecting antibody responses to Omicron, which would explain why binding Ab levels in VIOLA increased after booster but not after Omicron. Studies that fail to detect a rise in anti-Spike antibody levels following Omicron infection should be interpreted cautiously in this context of assays used.^44^ An increase in cellular response after infection in patients who have had two vaccine doses would be expected, given that ancestral T-cells efficiently cross-recognize the Omicron variant of SARS-CoV-2.^50^ It is less clear why cellular immunity appears to have declined after infections in patients with three vaccine doses. Perhaps the decline reflects T-cell exhaustion, as reported in infected cancer patients who had three doses of vaccine.^51^ However, T-cell exhaustion was not corroborated in a recent report in which triple-vaccinated, SARS-CoV-2-infected individuals had maximal frequency IFNγ expression to Spike protein.^52^ Reassuringly, VIOLA participants had stable COVID-19 v3 scores, following Omicron infection, as well as boosters (Fig. 7), and had relatively mild severity of infections: only 8% were hospitalized, and none had respiratory failure.

Among the strengths of VIOLA is that samples were collected from each patient at multiple strictly defined post-vaccine time points up to 2 years from the primary vaccination, allowing for rigorous assessment of the longitudinal trajectory of post-primary and post-booster vaccine responses. The collection of pre-vaccine blood samples enabled us to assess for serologic evidence of pre-existing COVID-19 exposure and to measure the magnitude of changes induced by the vaccination. We carefully monitored for any incident infections in order to disentangle the impact of vaccinations from that of infections. The two-center design allowed enrollment from the East Coast and Midwest locales with different race/ethnic compositions. Successful recruitment of under-represented minorities, who comprised 65% of the VIOLA cohort, led to a novel observation of stronger antibody responses among non-Whites. The diversity of immunoassays for measuring antibody and cellular responses allowed us to cross-check our conclusions with multiple tests and ensure that the results were not artifacts of specific methodology. Noteworthy as well is the use of live-virus neutralizing assays against Wuhan and Omicron variants, which is labor-intensive and requires a Biosafety level 3 laboratory, but makes possible a more granular analysis of antibody responses to different SARS-CoV-2 strains and vaccinations.

The main limitation of the study is the lack of longitudinal healthy controls. Instead, we analyzed sera from healthy controls from the NYU Vaccine Center and also included immunosequencing data from healthy subjects from the Adaptive dataset for reference. We also had to contend with missing data for participants who did not attend a scheduled study revisit or terminated the study early (Fig. 1). Because only 55% of the VIOLA participants opted for boosters, the statistical power of the ‘booster arm’ analyses was less than for primary series analyses. Lastly, although we assessed both humoral and cellular responses using a variety of assays, not all aspects of vaccine-induced immunity were examined. For example, we did not check for myeloid responses^53^ or for Spike-specific memory B-cells, which are challenging to detect in B-cell-depleted individuals.^25^

In conclusion, VIOLA demonstrates the serologic and cellular immune benefits of COVID-19 mRNA vaccines in B-cell-depleted MS patients, including in those with prior infection. Our results reinforce the importance of boosters, which induced serologic and cellular responses that were maintained up to 1-year after booster dose. Thus, annual COVID-19 vaccine dosing may be adequate in B-cell-depleted individuals, assuming that COVID-19 vaccines continue to be effective for the emergent SARS-CoV-2 variants. Future work should examine the immune impact of the new generation COVID-19 vaccines and assess longer-term immune memory to SARS-CoV-2 variants in B-cell depleted population.

## ACKNOWLEDGMENTS

The authors gratefully acknowledge our medical assistants, Teodora Drakulovic, April Oneal and Damaris Cloud, for their gentle bedside manner and great phlebotomy skills; our colleagues at NYUMCCC and RMMSC who assisted us with patient recruitment; and all the patients who participated in this study. Editorial assistance with tables and figures was provided by Emma Bouck, PhD, of Nucleus Global and funded by Genentech, Inc.

## AUTHOR CONTRIBUTIONS

I. Kister, R. Curtin, A.L. Piquet, J. Pei, M.I. Samanovic, M.J. Mulligan, Y. Patskovsky, J. Priest, M. Cabatingan, R.C. Winger, M. Krogsgaard and G.J. Silverman contributed to the conception and design of the study. I. Kister, R. Curtin, A.L. Piquet, T. Borko, J. Pei, B.L. Banbury, T.E. Bacon, A. Kim, M. Tuen, Y. Velmurugu, S. Nyovanie, S. Selva, M.I. Samanovic, M.J. Mulligan, Y. Patskovsky, J. Priest, M. Cabatingan, R.C. Winger, M. Krogsgaard and G.J. Silverman contributed to the acquisition and analysis of data. I. Kister, R. Curtin, A.L. Piquet, T. Borko, J. Pei, B.L. Banbury, T.E. Bacon, A. Kim, M. Tuen, Y. Velmurugu, S. Nyovanie, S. Selva, M.I. Samanovic, M.J. Mulligan, Y. Patskovsky, J. Priest, M. Cabatingan, R.C. Winger, M. Krogsgaard and G.J. Silverman contributed to drafting the text or preparing the figures.

## POTENTIAL CONFLICTS OF INTEREST

This work was supported by an unrestricted investigator-initiated grant from Genentech. **IK** served on the scientific advisory board for Biogen Idec, Genentech, Alexion, EMDSerono; received consulting fees from Roche; and received research support from Guthy-Jackson Charitable Foundation, National Multiple Sclerosis Society, Biogen Idec, Serono, Genzyme, and Genentech/Roche; he receives royalties from Wolters Kluwer for ’Top 100 Diagnosis in Neurology’ (co-written with Jose Biller). **GJS** received honoraria from BMS, Eli Lilly and Human Biosciences, and research support from BMS, Genentech, Biogen, Lupus Research Alliance, NIH-NIAMS, NIH-NIAID and the Colton Center at NYU. **MK** is on the scientific advisory board for Merck, NexImmune and Genentech and received research support from Merck Sharp & Dohme Corp., a subsidiary of Merck & Co., Inc., Genentech, the Mark Foundation, NIH-NIGMS and NIH-NCI. **BLB** receives salary and owns stock from Adaptive Biotechnologies. **MJM** reported potential competing interests: laboratory research and clinical trials contracts for vaccines or MAB vs SARS-CoV-2 with Lilly, Pfizer, and Sanofi; research grant funding from USG/HHS/NIH for vaccine and MAB clinical trials; personal fees for Scientific Advisory Board service from Merck, Meissa Vaccines, Inc. and Pfizer. **ALP** reports research grants from the University of Colorado, Rocky Mountain MS Center, and the Foundation for Sarcoidosis; consulting fees from Genentech/Roche, UCB, EMD Serono and Alexion; and honorarium from MedLink and publication royalties from Springer as co-editor of a medical textbook. All others declare no conflicts of interest.

## DATA AVAILABILITY

Anonymized data are available in **Table S2**.

## Notes

### Author Declarations

IRB of NYU Grossman School of Medicine (New York) has approved this work

## REFERENCES

1. Feng S, Phillips DJ, White T, et al. Correlates of protection against symptomatic and asymptomatic SARS-CoV-2 infection. Nat Med. 2021 Nov;27(11):2032–40.

2. Yamamoto S, Mizoue T, Ohmagari N. Analysis of Previous Infection, Vaccinations, and Anti-SARS-CoV-2 Antibody Titers and Protection Against Infection With the SARS-CoV-2 Omicron BA.5 Variant. JAMA Netw Open. 2023 Mar 1;6(3):e233370.

3. Kreuzberger N, Hirsch C, Andreas M, et al. Immunity after COVID-19 vaccination in people with higher risk of compromised immune status: a scoping review. Cochrane Database Syst Rev. 2022 Aug 9;8(8):CD015021.

4. Garjani A, Patel S, Bharkhada D, et al. Impact of mass vaccination on SARS-CoV-2 infections among multiple sclerosis patients taking immunomodulatory disease-modifying therapies in England. Mult Scler Relat Disord. 2022 Jan;57:103458.

5. Schiavetti I, Cordioli C, Stromillo ML, et al. Breakthrough SARS-CoV-2 infections in MS patients on disease-modifying therapies. Mult Scler. 2022 Nov;28(13):2106–11.

6. Jakimovski D, Zakalik K, Awan S, et al. COVID-19 Vaccination in Multiple Sclerosis and Inflammatory Diseases: Effects from Disease-Modifying Therapy, Long-Term Seroprevalence and Breakthrough Infections. Vaccines (Basel). 2022 Apr 28;10(5).

7. Zaloum SA, Wood CH, Tank P, et al. Risk of COVID-19 in people with multiple sclerosis who are seronegative following vaccination. Mult Scler. 2023 Jul;29(8):979–89.

8. Smith JB, Gonzales EG, Li BH, Langer-Gould A. Analysis of Rituximab Use, Time Between Rituximab and SARS-CoV-2 Vaccination, and COVID-19 Hospitalization or Death in Patients With Multiple Sclerosis. JAMA Netw Open. 2022 Dec 1;5(12):e2248664.

9. Liu N, Yu W, Sun M, et al. Outcome of COVID-19 Infection in Patients With Multiple Sclerosis Who Received Disease-Modifying Therapies: A Systematic Review and Meta-Analysis. J Clin Neurol. 2023 Jul;19(4):381–91.

10. Weberpals J, Roumpanis S, Barer Y, et al. Clinical outcomes of COVID-19 in patients with multiple sclerosis treated with ocrelizumab in the pre- and post-SARS-CoV-2 vaccination periods: Insights from Israel. Mult Scler Relat Disord. 2022 Dec;68:104153.

11. Sormani MP, Inglese M, Schiavetti I, et al. Effect of SARS-CoV-2 mRNA vaccination in MS patients treated with disease modifying therapies. EBioMedicine. 2021 Oct;72:103581.

12. Thompson AJ, Banwell BL, Barkhof F, et al. Diagnosis of multiple sclerosis: 2017 revisions of the McDonald criteria. Lancet Neurol. 2018 Feb;17(2):162–73.

13. Kister I, Patskovsky Y, Curtin R, et al. Cellular and Humoral Immunity to SARS-CoV-2 Infection in Multiple Sclerosis Patients on Ocrelizumab and Other Disease-Modifying Therapies: A Multi-Ethnic Observational Study. Ann Neurol. 2022 Jun;91(6):782–95.

14. CDC. CDC booster. Available from: https://www.cdc.gov/media/releases/2022/s0519-covid-booster-acip.html.

15. Radke EE, Brown SM, Pelzek AJ, et al. Hierarchy of human IgG recognition within the Staphylococcus aureus immunome. Sci Rep. 2018 Sep 5;8(1):13296.

16. Samanovic MI, Cornelius AR, Gray-Gaillard SL, et al. Robust immune responses are observed after one dose of BNT162b2 mRNA vaccine dose in SARS-CoV-2-experienced individuals. Sci Transl Med. 2022 Feb 9;14(631):eabi8961.

17. Elyanow R, Snyder TM, Dalai SC, et al. T cell receptor sequencing identifies prior SARS-CoV-2 infection and correlates with neutralizing antibodies and disease severity. JCI Insight. 2022 May 23;7(10).

18. Snyder TM, Gittelman RM, Klinger M, et al. Magnitude and Dynamics of the T-Cell Response to SARS-CoV-2 Infection at Both Individual and Population Levels. medRxiv. 2020 Sep 17.

19. Kister I, Curtin R, Pei J, et al. Hybrid and vaccine-induced immunity against SAR-CoV-2 in MS patients on different disease-modifying therapies. Ann Clin Transl Neurol. 2022 Oct;9(10):1643–59.

20. Tsang NNY, So HC, Cowling BJ, Leung GM, Ip DKM. Effectiveness of BNT162b2 and CoronaVac COVID-19 vaccination against asymptomatic and symptomatic infection of SARS-CoV-2 omicron BA.2 in Hong Kong: a prospective cohort study. Lancet Infect Dis. 2023 Apr;23(4):421–34.

21. Altarawneh HN, Chemaitelly H, Ayoub H, et al. Effect of prior infection, vaccination, and hybrid immunity against symptomatic BA.1 and BA.2 Omicron infections and severe COVID-19 in Qatar. 2022 2022-03-22.

22. Louapre C, Belin L, Marot S, et al. Three to four mRNA COVID-19 vaccines in multiple sclerosis patients on immunosuppressive drugs: Seroconversion and variant neutralization. Eur J Neurol. 2023 Sep;30(9):2781–92.

23. van Dam KPJ, Hogenboom L, Stalman EW, et al. Longitudinal SARS-CoV-2 humoral response in MS patients with and without SARS-CoV-2 infection prior to vaccination. Front Neurol. 2022 11/10/2022;13:1032830.

24. Sabatino J. Longitudinal adaptive immune responses following sequential SARS-CoV-2 vaccinations in MS patients on anti-CD20 therapies and sphingosine-1-phosphate receptor modulators. Multiple Sclerosis and Related Disorders. 2022.

25. Disanto G, Galante A, Cantu’ M, et al. Longitudinal Postvaccine SARS-CoV-2 Immunoglobulin G Titers, Memory B-Cell Responses, and Risk of COVID-19 in Multiple Sclerosis Over 1 Year. 2023 2023-01-01.

26. Petrone L, Tortorella C, Aiello A, et al. Humoral and Cellular Response to Spike of Delta SARS-CoV-2 Variant in Vaccinated Patients With Multiple Sclerosis. Front Neurol. 2022 05/31/2022;13:881988.

27. Brill L, Rechtman A, Zveik O, et al. Humoral and T-Cell Response to SARS-CoV-2 Vaccination in Patients With Multiple Sclerosis Treated With Ocrelizumab. JAMA Neurol. 2021 Dec 1;78(12):1510–4.

28. Levin EG, Lustig Y, Cohen C, et al. Waning Immune Humoral Response to BNT162b2 Covid-19 Vaccine over 6 Months. N Engl J Med. 2021 Dec 9;385(24):e84.

29. Laidlaw BJ, Ellebedy AH. The germinal centre B cell response to SARS-CoV-2. Nat Rev Immunol. 2022 Jan;22(1):7–18.

30. Maglione A, Morra M, Meroni R, Matta M, Clerico M, Rolla S. Humoral response after the booster dose of anti-SARS-CoV-2 vaccine in multiple sclerosis patients treated with high-efficacy therapies. Mult Scler Relat Disord. 2022 May;61:103776.

31. Schiavetti I, Inglese M, Frau J, et al. Antibody response elicited by the SARS-CoV-2 vaccine booster in patients with multiple sclerosis: Who gains from it? Eur J Neurol. 2023 Aug;30(8):2357–64.

32. Achtnichts L, Jakopp B, Oberle M, et al. Humoral Immune Response after the Third SARS-CoV-2 mRNA Vaccination in CD20 Depleted People with Multiple Sclerosis. Vaccines (Basel). 2021 Dec 11;9(12).

33. Bajwa HM, Novak F, Nilsson AC, et al. Persistently reduced humoral and sustained cellular immune response from first to third SARS-CoV-2 mRNA vaccination in anti-CD20-treated multiple sclerosis patients. Mult Scler Relat Disord. 2022 Apr;60:103729.

34. Samanovic MI, Oom AL, Cornelius AR, et al. Vaccine-Acquired SARS-CoV-2 Immunity versus Infection-Acquired Immunity: A Comparison of Three COVID-19 Vaccines. Vaccines (Basel). 2022 Dec 15;10(12).

35. Branche AR, Rouphael NG, Diemert DJ, et al. Comparison of bivalent and monovalent SARS-CoV-2 variant vaccines: the phase 2 randomized open-label COVAIL trial. Nat Med. 2023 Sep;29(9):2334–46.

36. Thompson PW. Efficacy of Covid-19 Vaccines in Ethnically Diverse Population (BAME): A Systematic Review. Pakistan Journal of Medical & Health Sciences. 2022;16.

37. Kurupati R, Kossenkov A, Haut L, et al. Race-related differences in antibody responses to the inactivated influenza vaccine are linked to distinct pre-vaccination gene expression profiles in blood. Oncotarget. 2016 Sep 27;7(39):62898–911.

38. Haralambieva IH, Salk HM, Lambert ND, et al. Associations between race, sex and immune response variations to rubella vaccination in two independent cohorts. Vaccine. 2014 Apr 7;32(17):1946–53.

39. Saidenberg L, Arbini AA, Silverman GJ, Lotan I, Cutter G, Kister I. Faster B-cell repletion after anti-CD20 infusion in Black patients compared to white patients with neurologic diseases. Mult Scler Relat Disord. 2022 Jul;63:103830.

40. Rauber S, Korsen M, Huntemann N, et al. Immune response to SARS-CoV-2 vaccination in relation to peripheral immune cell profiles among patients with multiple sclerosis receiving ocrelizumab. J Neurol Neurosurg Psychiatry. 2022 Feb 22.

41. Sabatino JJ, Jr., Mittl K, Rowles WM, et al. Multiple sclerosis therapies differentially affect SARS-CoV-2 vaccine-induced antibody and T cell immunity and function. JCI Insight. 2022 Feb 22;7(4).

42. Brill L, Raposo C, Rechtman A, et al. Severe Acute Respiratory Syndrome Coronavirus 2 Third Vaccine Immune Response in Multiple Sclerosis Patients Treated with Ocrelizumab. Ann Neurol. 2022 Jun;91(6):796–800.

43. Hvidt AK, Guo H, Andersen R, et al. Long-term humoral and cellular immunity after primary SARS-CoV-2 infection: a 20-month longitudinal study. BMC Immunology. 2023 2023-11-16;24(1):1–13.

44. Maglione A, Francese R, Arduino I, et al. Long-lasting neutralizing antibodies and T cell response after the third dose of mRNA anti-SARS-CoV-2 vaccine in multiple sclerosis. Front Immunol. 2023 06/19/2023;14:1205879.

45. Ruggieri S, Aiello A, Tortorella C, et al. Dynamic Evolution of Humoral and T-Cell Specific Immune Response to COVID-19 mRNA Vaccine in Patients with Multiple Sclerosis Followed until the Booster Dose. Int J Mol Sci. 2023 May 10;24(10).

46. Aiello A, Coppola A, Ruggieri S, et al. Longitudinal characterisation of B and T-cell immune responses after the booster dose of COVID-19 mRNA-vaccine in people with multiple sclerosis using different disease-modifying therapies. J Neurol Neurosurg Psychiatry. 2023 Apr;94(4):290–9.

47. Palomares Cabeza V, Kummer LYL, Wieske L, et al. Longitudinal T-Cell Responses After a Third SARS-CoV-2 Vaccination in Patients With Multiple Sclerosis on Ocrelizumab or Fingolimod. Neurol Neuroimmunol Neuroinflamm. 2022 Jul;9(4).

48. Madelon N, Heikkila N, Sabater Royo I, et al. Omicron-Specific Cytotoxic T-Cell Responses After a Third Dose of mRNA COVID-19 Vaccine Among Patients With Multiple Sclerosis Treated With Ocrelizumab. JAMA Neurol. 2022 Apr 1;79(4):399–404.

49. Algu P, Hameed N, DeAngelis T, Stern J, Harel A. Post-vaccination SARS-Cov-2 T-cell receptor repertoires in patients with multiple sclerosis and related disorders. Mult Scler Relat Disord. 2023 Nov;79:104965.

50. Tarke A, Sidney J, Methot N, et al. Impact of SARS-CoV-2 variants on the total CD4(+) and CD8(+) T cell reactivity in infected or vaccinated individuals. Cell Rep Med. 2021 Jul 20;2(7):100355.

51. Benitez Fuentes JD, Mohamed Mohamed K, de Luna Aguilar A, et al. Evidence of exhausted lymphocytes after the third anti-SARS-CoV-2 vaccine dose in cancer patients. Front Oncol. 2022 12/20/2022;12:975980.

52. Cai C, Gao Y, Adamo S, et al. SARS-CoV-2 vaccination enhances the effector qualities of spike-specific T cells induced by COVID-19. Sci Immunol. 2023 Dec 8;8(90):eadh0687.

53. Wang M, Dehlinger A, Zapata CF, et al. Associations of myeloid cells with cellular and humoral responses following vaccinations in patients with neuroimmunological diseases. Nat Commun. 2023 Nov 25;14(1):7728.

