## Supplemental Table S1 and Figure S1 for "Longitudinal study of immunity to SARS-CoV2 in Ocrelizumab-treated multiple sclerosis patients up to 2 years after COVID-19 vaccination"

**Table S1. Demographics and clinical characteristics of participants who did or did not receive a COVID-19 booster**

| **Characteristic** | **All VIOLA**  **N=60** | **Booster**  **n=33** | **No Booster**  **n=27** | ***P* Value^a^** |
| --- | --- | --- | --- | --- |
| **Age, years** |  |  |  |  |
| Mean (SD) | 38.6 (10.0) | 38.3 (10.3) | 38.9 (9.8) | 0.8 |
| Median (range) | 37.0 (20.0–57.7) | 36.4 (20.0–57.7) | 37.2 (21.1–55.2) |  |
| **Female, n (%)** | 44 (73.3) | 25 (75.8) | 19 (70.4) | 0.8 |
| **BMI, mean (SD), kg/m^2^** | 31.0 (9.0) | 30.0 (8.8) | 32.2 (9.3) | 0.4 |
| **Race and ethnicity, n (%)** |  |  |  |  |
| African American/Black | 16 (26.7) | 8 (24.2) | 8 (29.6) | 0.3 |
| Hispanic/Latino | 13 (21.7) | 8 (24.4) | 5 (18.5) |  |
| White | 27 (45.0) | 13 (39.4) | 14 (51.9) |  |
| Other | 4 (6.7) | 4 (12.1) | 0 (0) |  |
| **MS subtype, n (%)** |  |  |  |  |
| Relapsing-remitting | 56 (93.3) | 30 (90.9) | 26 (96.3) | 0.9 |
| Primary progressive | 0 (0) | 0 (0) | 0 (0) |  |
| Secondary progressive | 3 (5.0) | 3 (9.1) | 0 (0) |  |
| Primary relapsing | 1 (1.7) | 0 (0) | 1 (3.7) |  |
| **EDSS score** |  |  |  |  |
| Mean (SD) | 2.4 (1.8) | 2.3 (1.9) | 2.6 (1.7) | 0.6 |
| Median (range) | 2.0 (0–6.5) | 2.0 (0–6.5) | 2.5 (0–6.5) |  |
| **Time since onset of MS symptoms, mean (SD), weeks^b^** | 429.8 (320.2) | 411.0 (366.3) | 452.8 (257.8) | 0.6 |
| **Duration of OCR treatment, mean (SD), weeks^b^** | 88.1 (58.6) | 78.1 (55.1) | 100.3 (61.5) | 0.2 |
| **Time from last OCR infusion to 2^nd^ vaccine dose, mean (SD), weeks** | 19.0 (6.4) | 19.5 (5.9) | 18.4 (7.0) | 0.5 |
| **Number of COVID-relevant comorbidities, n (%)^c^** |  |  |  |  |
| 0 | 23 (38.3) | 13 (39.4) | 10 (37.0) | 0.9 |
| 1 | 17 (28.3) | 9 (27.3) | 8 (29.6) |  |
| 2 | 12 (20.0) | 7 (21.2) | 5 (18.5) |  |
| ≥3 | 8 (13.3) | 4 (12.2) | 4 (14.8) |  |
| **COVID-19 diagnosis prior to vaccination, n (%)** | 27 (45.0) | 15 (45.5) | 12 (44.4) | 1 |
| **Vaccination type, n (%)** |  |  |  |  |
| Moderna | 11 (18.3) | 3 (9.1) | 8 (29.6) | 0.05 |
| Pfizer | 49 (81.7) | 30 (90.9) | 19 (70.4) |  |

BMI, body mass index; EDSS, Expanded Disability Status Scale; MS, multiple sclerosis; NYU, New York University; OCR, ocrelizumab; CU-AMC, University of Colorado Anschutz Medical Center.

^a^*P* values compare Booster and No Booster subgroups and were determined via two-group t-test for continuous and quantitative variables, via Mann-Whitney U-test for ordinal variables and via Fisher’s exact test for categorical variables.

^b^The reported durations ended at last OCR infusion before enrolling in the study.

^c”^COVID-relevant comorbidities” included hypertension, chronic obstructive pulmonary disease, cardiovascular disease, diabetes mellitus, sickle cell disease, chronic kidney disease, chronic liver disease and (non-skin) cancer.

**Figure S1: Timeline of vaccinations, infections and OCR infusions relative to sample collection times for the 16 patients with immunosequencing data**

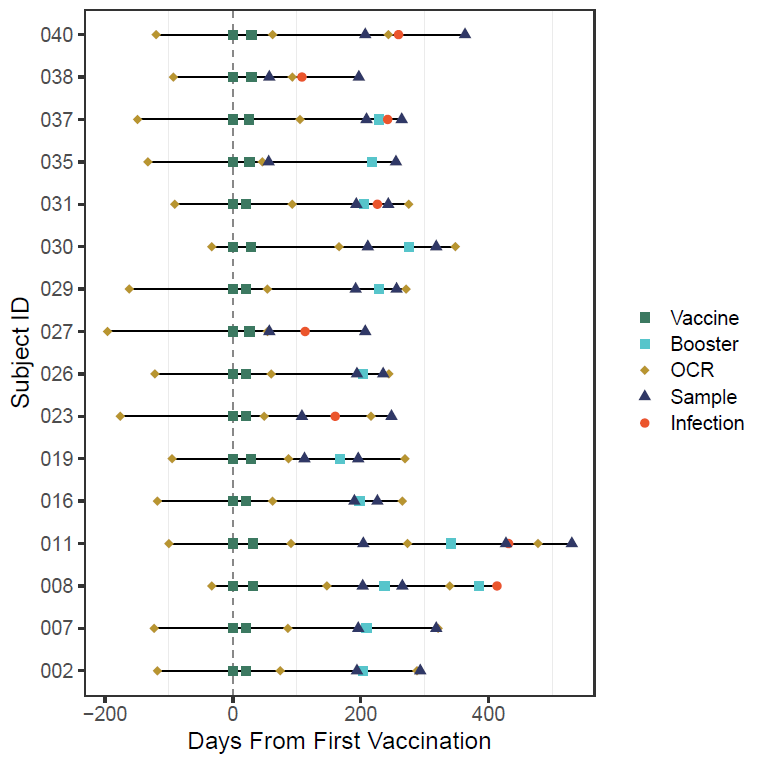

OCR, ocrelizumab.

The first two doses of vaccines are shown as dark green squares, and the third dose as turquoise squares.
