## Supplemental Table S2 for "Longitudinal study of immunity to SARS-CoV2 in Ocrelizumab-treated multiple sclerosis patients up to 2 years after COVID-19 vaccination"

| annonymizedid | redcap_event_name | assay_sample_date_collecte | anti_sars_cov_2_s_quant | t_cell_elispot_ifng | t_cell_elispot_il_2 | igg_spike_1_1000_igg_spike | covid_count | monoclonals_yn | ic50WT | ic50Omicron |
|---|---|---|---|---|---|---|---|---|---|---|
| 40 | baseline_arm_1 | 4/22/2021 | 0.4 | 30 | 0 | 279 | 1 | 0 | | |
| 40 | 4_weeks_post_dose_arm_1 | 7/15/2021 | 0.4 | 170 | 90 | 39 | 1 | 0 | 10 | 10 |
| 40 | 12_weeks_post_dose_arm_1 | 9/8/2021 | 0.4 | 390 | 200 | 47 | 1 | 0 | | |
| 40 | 24_weeks_post_dose_arm_1 | 12/7/2021 | 0.4 | 250 | 50 | 260 | 1 | 0 | | |
| 40 | bv_4_weeks_post_do_arm_2 | 1/14/2022 | 0.4 | No PBMC | No PBMC | 260 | 1 | 0 | | |
| 40 | bv_12_weeks_post_d_arm_2 | 3/16/2022 | 0.4 | 1880 | 600 | 193 | 1 | 0 | 10 | 10 |
| 40 | bv_24_weeks_post_d_arm_2 | 6/8/2022 | 9904 | | | 834 | 1 | 1 | 10 | 10 |
| 40 | bv_48_weeks_post_d_arm_2 | 11/16/2022 | 897 | 210 | 40 | | 1 | 1 | | |
| 28 | baseline_arm_1 | 4/26/2021 | 9.43 | 220 | 160 | 3467 | 1 | 0 | 27 | 10 |
| 28 | 4_weeks_post_dose_arm_1 | 7/16/2021 | 387.3 | PBMC LOST | PBMC LOST | 12406 | 1 | 0 | 177 | 10 |
| 28 | 12_weeks_post_dose_arm_1 | 9/10/2021 | 106 | 410 | 0 | 2370 | 1 | 0 | | |
| 28 | 24_weeks_post_dose_arm_1 | 12/2/2021 | 56.97 | 430 | 10 | 2041 | 1 | 0 | | |
| 28 | 48_weeks_post_dose_arm_1 | 6/3/2022 | 21.5 | | | 10185.25 | 1 | 0 | | |
| 57 | baseline_arm_1 | 4/29/2021 | 0.4 | 10 | 10 | 1704 | 1 | 0 | 10 | 10 |
| 57 | 4_weeks_post_dose_arm_1 | 7/16/2021 | 0.4 | PBMC LOST | PBMC LOST | 5971 | 2 | 0 | 10 | 10 |
| 57 | 12_weeks_post_dose_arm_1 | 9/10/2021 | 0.95 | 1660 | 80 | 2534 | 2 | 0 | | |
| 57 | 24_weeks_post_dose_arm_1 | 12/2/2021 | 0.4 | 190 | 10 | 1629 | 2 | 0 | | |
| 57 | bv_4_weeks_post_do_arm_2 | 3/31/2022 | 0.4 | 3780 | 580 | 15864.75 | 2 | 0 | 10 | 10 |
| 57 | bv_12_weeks_post_d_arm_2 | 5/26/2022 | 0.5 | 6000 | 1820 | 13935.25 | 2 | 0 | | |
| 57 | bv_24_weeks_post_d_arm_2 | 8/30/2022 | 0.57 | 3890 | 270 | 12005.25 | 2 | 0 | 10 | 10 |
| 57 | bv_48_weeks_post_d_arm_2 | 2/21/2023 | 0.43 | 7150 | 20 | 11901.25 | 2 | 0 | | |
| 7 | baseline_arm_1 | 1/25/2021 | 0.4 | No PBMC | No PBMC | 43 | 0 | 0 | 10 | 10 |
| 7 | 4_weeks_post_dose_arm_1 | 5/19/2021 | 4725 | 490 | 830 | 29653 | 0 | 0 | 5851.5 | 26.18 |
| 7 | 12_weeks_post_dose_arm_1 | 7/14/2021 | 3966 | PBMC LOST | PBMC LOST | 28055 | 0 | 0 | | |
| 41 | baseline_arm_1 | 1/15/2021 | 0.4 | No response | No response | 192 | 1 | 0 | 10 | |
| 41 | 4_weeks_post_dose_arm_1 | 7/19/2021 | 0.4 | PBMC LOST | PBMC LOST | 160 | 1 | 0 | 14 | 10 |
| 41 | 12_weeks_post_dose_arm_1 | 9/13/2021 | 0.4 | 40 | 50 | 170 | 1 | 0 | | |
| 41 | 24_weeks_post_dose_arm_1 | 12/6/2021 | 0.4 | 50 | 0 | 248 | 1 | 0 | | |
| 41 | bv_24_weeks_post_d_arm_2 | 6/7/2022 | 0.4 | 200 | 20 | 993.5 | 1 | 0 | 10 | 10 |
| 41 | bv_48_weeks_post_d_arm_2 | 11/21/2022 | 0.4 | 450 | 20 | 2765 | 2 | 0 | | |
| 30 | baseline_arm_1 | 3/24/2021 | 0.4 | 23 | 7 | 537 | 1 | 0 | 10 | 10 |
| 30 | 4_weeks_post_dose_arm_1 | 6/11/2021 | 0.4 | 220 | 210 | 53 | 1 | 0 | 16 | 10 |
| 30 | 12_weeks_post_dose_arm_1 | 8/3/2021 | 0.4 | PBMC LOST | PBMC LOST | 47 | 1 | 0 | | |
| 30 | 24_weeks_post_dose_arm_1 | 10/29/2021 | 0.4 | 100 | 40 | 200 | 1 | 0 | | |
| 30 | bv_48_weeks_post_d_arm_2 | 11/21/2021 | 241 | No PHA | No PHA | 13799 | 1 | 0 | | |

| | | | | | | | | | | |
|---|---|---|---|---|---|---|---|---|---|---|
| 30 | bv_4_weeks_post_do_arm_2 | 12/9/2021 | 0.4 | No PBMC | No PBMC | 2173.75 | 1 | 0 | 10 | 10 |
| 30 | bv_12_weeks_post_d_arm_2 | 2/28/2022 | 0.4 | 130 | 160 | 1829.75 | 1 | 0 | 10 | 10 |
| 30 | bv_24_weeks_post_d_arm_2 | 5/2/2022 | 3394 | 350 | 110 | 15505.25 | 1 | 1 | 55622 | 16.02333 |
| 52 | baseline_arm_1 | 2/5/2021 | 0.4 | 15 | 0 | 168 | 1 | 0 | 10 | 10 |
| 52 | 4_weeks_post_dose_arm_1 | 5/13/2021 | 0.4 | 247 | 43 | 41 | 1 | 0 | 14 | 10 |
| 52 | 12_weeks_post_dose_arm_1 | 7/8/2021 | 0.4 | PBMC LOST | PBMC LOST | 30 | 1 | 0 | | |
| 52 | 24_weeks_post_dose_arm_1 | 10/5/2021 | 0.4 | 20 | 0 | 41 | 1 | 0 | | |
| 52 | bv_4_weeks_post_do_arm_2 | 12/6/2021 | 0.4 | 80 | 20 | 206 | 1 | 0 | 10 | 10 |
| 52 | bv_12_weeks_post_d_arm_2 | 2/24/2022 | 0.4 | 140 | 250 | 18.75 | 1 | 0 | | |
| 52 | bv_48_weeks_post_d_arm_2 | 10/24/2022 | 471.8 | 110 | 120 | | 2 | 1 | | |
| 29 | baseline_arm_1 | 3/10/2021 | 0.4 | 990 | 473 | 113 | 0 | 0 | 48.66 | 10 |
| 29 | 4_weeks_post_dose_arm_1 | 5/27/2021 | 14.3 | 597 | 173 | 6220 | 0 | 0 | 25 | 10 |
| 29 | 12_weeks_post_dose_arm_1 | 7/22/2021 | 8.43 | PBMC LOST | PBMC LOST | 2070 | 0 | 0 | | |
| 29 | 24_weeks_post_dose_arm_1 | 10/15/2021 | 6.2 | 60 | 0 | 926 | 0 | 0 | | |
| 29 | bv_4_weeks_post_do_arm_2 | 12/1/2021 | 7568 | 440 | 110 | 11404.75 | 0 | 0 | 582 | 38.275 |
| 29 | bv_12_weeks_post_d_arm_2 | 2/18/2022 | 14547 | 250 | 100 | 18521.75 | 1 | 1 | | |
| 29 | bv_24_weeks_post_d_arm_2 | 4/21/2022 | 9390 | 190 | 10 | 20689 | 1 | 1 | 619.5 | 167.6 |
| 58 | baseline_arm_1 | 1/27/2021 | 0.4 | No response | No response | 132 | 0 | 0 | 10 | 10 |
| 58 | 4_weeks_post_dose_arm_1 | 5/14/2021 | 1.04 | 443 | 130 | 1651 | 0 | 0 | 10 | 10 |
| 58 | 12_weeks_post_dose_arm_1 | 7/19/2021 | 0.96 | PBMC LOST | PBMC LOST | 14131 | 0 | 0 | | |
| 58 | 24_weeks_post_dose_arm_1 | 10/7/2021 | 0.75 | No PHA | No PHA | 297 | 0 | 0 | | |
| 58 | bv_4_weeks_post_do_arm_2 | 3/16/2022 | 1.51 | 450 | 100 | 9988.75 | 1 | 0 | 10 | 10 |
| 58 | bv_12_weeks_post_d_arm_2 | 5/9/2022 | 1.25 | 710 | 350 | 11853.5 | 1 | 0 | | |
| 58 | bv_24_weeks_post_d_arm_2 | 7/26/2022 | 185.3 | 230 | 70 | 13227 | 2 | 1 | 3975.5 | 5883 |
| 58 | bv_48_weeks_post_d_arm_2 | 2/6/2023 | 1.68 | 690 | 500 | 5185.25 | 2 | 1 | | |
| 59 | baseline_arm_1 | 2/16/2021 | 0.4 | No response | No response | 176 | 1 | 0 | 10 | 10 |
| 59 | 4_weeks_post_dose_arm_1 | 5/17/2021 | 3.89 | 220 | 103 | 8304 | 1 | 0 | 10 | 10 |
| 59 | 12_weeks_post_dose_arm_1 | 7/26/2021 | 2.05 | PBMC LOST | PBMC LOST | 1663 | 1 | 0 | | |
| 59 | 24_weeks_post_dose_arm_1 | 9/30/2021 | 1.05 | 0 | 0 | 158 | 1 | 0 | | |
| 59 | bv_12_weeks_post_d_arm_2 | 5/11/2022 | 0.52 | 310 | 770 | 3895 | 1 | 0 | | |
| 59 | bv_24_weeks_post_d_arm_2 | 8/22/2022 | 9.05 | 30 | 40 | 6563.25 | 2 | 0 | 10 | 10 |
| 59 | bv_48_weeks_post_d_arm_2 | 1/26/2023 | 2.75 | 40 | 140 | 9883 | 2 | 0 | | |
| 46 | baseline_arm_1 | 5/18/2021 | 0.4 | 0 | 0 | 134 | 0 | 0 | 13 | 10 |
| 46 | 4_weeks_post_dose_arm_1 | 8/25/2021 | 4.23 | 50 | 20 | 3411 | 0 | 0 | 114 | 27.88 |
| 46 | 12_weeks_post_dose_arm_1 | 10/21/2021 | 14.6 | 10 | 0 | 2525 | 0 | 0 | | |
| 46 | 24_weeks_post_dose_arm_1 | 1/11/2022 | 11.26 | 850 | 440 | 1873 | 0 | 0 | | |
| 46 | bv_4_weeks_post_do_arm_2 | 2/24/2022 | 3514 | 250 | 30 | 21456.75 | 1 | 0 | 1726.5 | 15.33333 |
| 46 | bv_12_weeks_post_d_arm_2 | 4/27/2022 | 170.5 | 2160 | 2010 | 3374 | 1 | 0 | 478.25 | 22 |
| 46 | bv_24_weeks_post_d_arm_2 | 7/19/2022 | 137.9 | 690 | 540 | 15169 | 1 | 0 | 77 | 10 |

| | | | | | | | | | | |
|---|---|---|---|---|---|---|---|---|---|---|
| 46 | bv_48_weeks_post_d_arm_2 | 1/9/2023 | 148.9 | 160 | 160 | 15615.5 | 2 | 0 | | |
| 54 | baseline_arm_1 | 5/19/2021 | 0.4 | 190 | 80 | 1095 | 1 | 0 | 17 | 10 |
| 54 | 4_weeks_post_dose_arm_1 | 7/13/2021 | 4949 | No PHA | No PHA | 23288 | 1 | 0 | 1351.033 | 10 |
| 54 | 12_weeks_post_dose_arm_1 | 8/31/2021 | 2974 | 2780 | 150 | 17913 | 1 | 0 | | |
| 54 | 24_weeks_post_dose_arm_1 | 11/30/2021 | 1560 | 260 | 20 | 9045 | 1 | 0 | | |
| 54 | 48_weeks_post_dose_arm_1 | 5/12/2022 | 694 | No PHA | No PHA | 7990 | 1 | 0 | 68 | 21.08 |
| 54 | bv_4_weeks_post_do_arm_2 | 8/19/2022 | 810 | 2130 | 20 | 12063.25 | 1 | 0 | 88 | 10 |
| 54 | bv_12_weeks_post_d_arm_2 | 10/19/2022 | | 3940 | 140 | 9913.5 | 1 | 0 | | |
| 54 | bv_24_weeks_post_d_arm_2 | 1/10/2023 | 534.5 | 7260 | 1130 | 11044 | 1 | 0 | | |
| 20 | baseline_arm_1 | 1/15/2021 | 0.4 | 0 | 7 | 17 | 0 | 0 | | |
| 20 | 4_weeks_post_dose_arm_1 | 9/2/2021 | 0.4 | 300 | 50 | 600 | 0 | 0 | 10 | 10 |
| 20 | 12_weeks_post_dose_arm_1 | 11/4/2021 | 92.5 | No PHA | No PHA | 23004 | 1 | 1 | | |
| 20 | 24_weeks_post_dose_arm_1 | 1/31/2022 | 1053 | 1490 | 860 | 24738 | 1 | 1 | | |
| 20 | 48_weeks_post_dose_arm_1 | 7/11/2022 | 30.4 | 500 | 150 | 16072.25 | 1 | 1 | 42.75 | 10 |
| 24 | baseline_arm_1 | 4/1/2021 | 0.4 | 140 | 30 | 200 | 1 | 0 | 10 | 10 |
| 24 | 4_weeks_post_dose_arm_1 | 5/25/2021 | 0.4 | 460 | 180 | 6301 | 1 | 0 | 10 | 10 |
| 24 | 12_weeks_post_dose_arm_1 | 7/21/2021 | 0.4 | PBMC LOST | PBMC LOST | 2675 | 1 | 0 | | |
| 24 | 24_weeks_post_dose_arm_1 | 10/11/2021 | 0.4 | 40 | 0 | 779 | 1 | 0 | | |
| 24 | 48_weeks_post_dose_arm_1 | 4/6/2022 | 0.4 | 110 | 0 | 4190.25 | 2 | 0 | 16 | 39.34667 |
| 44 | baseline_arm_1 | 5/27/2021 | 0.4 | 20 | 100 | 152 | 0 | 0 | 11 | 10 |
| 44 | 4_weeks_post_dose_arm_1 | 8/10/2021 | 13.84 | 10 | 30 | 3250 | 0 | 0 | 10 | 10 |
| 44 | 12_weeks_post_dose_arm_1 | 9/30/2021 | 5.01 | 10 | 0 | 1704 | 0 | 0 | | |
| 44 | 24_weeks_post_dose_arm_1 | 12/22/2021 | 16 | 40 | 20 | 455 | 0 | 0 | | |
| 44 | bv_4_weeks_post_do_arm_2 | 1/27/2022 | 1239 | 190 | 140 | 19668.5 | 0 | 0 | 89 | 24.67333 |
| 44 | bv_12_weeks_post_d_arm_2 | 3/24/2022 | 243.2 | 220 | 90 | 17624.5 | 0 | 0 | 30 | 10 |
| 44 | bv_24_weeks_post_d_arm_2 | 6/15/2022 | 66.4 | 20 | 70 | 11626.75 | 1 | 0 | 10 | 35.21667 |
| 44 | bv_48_weeks_post_d_arm_2 | 12/13/2022 | 22.8 | 250 | 100 | 10077.75 | 1 | 0 | | |
| 43 | baseline_arm_1 | 4/6/2021 | 0.4 | No response | No response | 114 | 0 | 0 | 16 | 10 |
| 43 | 4_weeks_post_dose_arm_1 | 5/27/2021 | 24.99 | 197 | 87 | 4732 | 0 | 0 | 10 | 10 |
| 43 | 12_weeks_post_dose_arm_1 | 7/26/2021 | 6.47 | PBMC LOST | PBMC LOST | 2411 | 0 | 0 | | |
| 43 | 24_weeks_post_dose_arm_1 | 10/19/2021 | 1.31 | 10 | 0 | 469 | 0 | 0 | | |
| 43 | bv_4_weeks_post_do_arm_2 | 11/22/2021 | 62.69 | 330 | 150 | 21294.75 | 0 | 0 | 67 | 10 |
| 43 | bv_12_weeks_post_d_arm_2 | 1/12/2022 | 11.04 | 100 | 220 | 19785.5 | 0 | 0 | | |
| 43 | bv_24_weeks_post_d_arm_2 | 4/11/2022 | 2.51 | 100 | 70 | 14809.25 | 0 | 0 | 18.46 | 10 |
| 43 | bv_48_weeks_post_d_arm_2 | 9/30/2022 | | 20 | 0 | | 0 | 0 | | |
| 27 | baseline_arm_1 | 3/29/2021 | 0.4 | 0 | 0 | 329 | 1 | 0 | 10 | 10 |
| 27 | 4_weeks_post_dose_arm_1 | 8/2/2021 | 44.63 | PBMC LOST | PBMC LOST | 17666 | 1 | 0 | 71.75 | 10 |
| 27 | 12_weeks_post_dose_arm_1 | 9/23/2021 | 150.6 | 180 | 10 | 15052 | 1 | 0 | | |
| 27 | 24_weeks_post_dose_arm_1 | 12/20/2021 | 38.7 | No PBMC | No PBMC | 6376.7 | 1 | 0 | | |

| | | | | | | | | | | |
|---|---|---|---|---|---|---|---|---|---|---|
| 27 | 48_weeks_post_dose_arm_1 | 6/15/2022 | 14.7 | 100 | 0 | 6620.5 | 1 | 0 | 10 | 10 |
| 47 | baseline_arm_1 | 2/9/2021 | 0.4 | 0 | 0 | 134 | 0 | 0 | 19 | 10 |
| 47 | 4_weeks_post_dose_arm_1 | 8/11/2021 | 463.9 | No PHA | No PHA | 5118 | 0 | 0 | 10 | 10 |
| 47 | 12_weeks_post_dose_arm_1 | 10/6/2021 | 69.1 | No PHA | No PHA | 5984 | 0 | 0 | | |
| 47 | bv_4_weeks_post_do_arm_2 | 12/29/2021 | 18.6 | 440 | 370 | 14503 | 0 | 0 | | |
| 47 | bv_12_weeks_post_d_arm_2 | 3/8/2022 | 8.76 | 840 | 400 | 14373.75 | 0 | 0 | 10 | 10 |
| 47 | bv_24_weeks_post_d_arm_2 | 6/7/2022 | 5.99 | 630 | 220 | 12115.5 | 0 | 0 | | |
| 47 | bv_48_weeks_post_d_arm_2 | 12/21/2022 | 4.8 | 1390 | 1030 | 15243 | 1 | 0 | | |
| 18 | baseline_arm_1 | 4/13/2021 | 0.4 | No PBMC | No PBMC | 147 | 0 | 0 | 10 | 10 |
| 18 | 4_weeks_post_dose_arm_1 | 6/8/2021 | 102.2 | 893 | 137 | 3583 | 0 | 0 | 10 | 10 |
| 18 | 12_weeks_post_dose_arm_1 | 8/6/2021 | 21.03 | 200 | 110 | 2697 | 0 | 0 | | |
| 18 | 24_weeks_post_dose_arm_1 | 11/4/2021 | 3.95 | 100 | 40 | 434 | 0 | 0 | | |
| 18 | bv_4_weeks_post_do_arm_2 | 1/25/2022 | 224 | 1960 | 660 | 17246.5 | 1 | 1 | 614.5 | 168 |
| 18 | bv_12_weeks_post_d_arm_2 | 3/28/2022 | 5755 | 4170 | 1560 | 15840.5 | 1 | 1 | | |
| 55 | baseline_arm_1 | 3/31/2021 | 0.4 | 0 | 13 | 293 | 1 | 0 | 10 | 10 |
| 55 | 4_weeks_post_dose_arm_1 | 6/8/2021 | 885.9 | 1040 | 380 | 4446 | 1 | 0 | 10 | 10 |
| 55 | 12_weeks_post_dose_arm_1 | 7/29/2021 | 181.8 | PBMC LOST | PBMC LOST | 1475 | 1 | 0 | | |
| 55 | 24_weeks_post_dose_arm_1 | 10/21/2021 | 44.7 | 410 | 180 | 338 | 1 | 0 | | |
| 55 | bv_4_weeks_post_do_arm_2 | 1/28/2022 | 213.2 | 610 | 60 | 32900.75 | 2 | 0 | | |
| 55 | bv_12_weeks_post_d_arm_2 | 3/30/2022 | 14059 | 1060 | 190 | 28555.25 | 2 | 0 | 10 | 62.58 |
| 55 | bv_24_weeks_post_d_arm_2 | 6/23/2022 | 3431 | 260 | 30 | 22020.5 | 2 | 0 | 10 | 10 |
| 55 | bv_48_weeks_post_d_arm_2 | 12/5/2022 | 1202 | 1150 | 520 | 23499.25 | 2 | 0 | | |
| 36 | baseline_arm_1 | 3/3/2021 | 0.4 | 0 | 0 | 421 | 1 | 0 | 10 | 10 |
| 36 | 4_weeks_post_dose_arm_1 | 7/8/2021 | 0.4 | 980 | 40 | 235 | 1 | 0 | 10 | 10 |
| 36 | bv_4_weeks_post_do_arm_2 | 9/14/2021 | 0.4 | 1800 | 760 | 13008.5 | 1 | 0 | 10 | 10 |
| 36 | bv_12_weeks_post_d_arm_2 | 11/19/2021 | 0.4 | 440 | 110 | 6580.75 | 1 | 0 | | |
| 36 | bv_24_weeks_post_d_arm_2 | 2/28/2022 | 0.4 | 80 | 0 | 13559.75 | 1 | 0 | 10 | 10 |
| 36 | bv_48_weeks_post_d_arm_2 | 7/26/2022 | 0.4 | 180 | 10 | 11459 | 2 | 0 | | |
| 19 | baseline_arm_1 | 3/25/2021 | 0.4 | 60 | 60 | 40 | 0 | 0 | 10 | 10 |
| 19 | 4_weeks_post_dose_arm_1 | 8/27/2021 | 632.8 | 240 | 220 | 10686 | 0 | 0 | 1412 | 24.61 |
| 19 | 12_weeks_post_dose_arm_1 | 10/22/2021 | 194 | 140 | 50 | 505 | 0 | 0 | | |
| 19 | 24_weeks_post_dose_arm_1 | 1/28/2022 | 9148 | 220 | 30 | 21917 | 1 | 1 | | |
| 19 | 48_weeks_post_dose_arm_1 | 7/1/2022 | 1741 | 330 | 210 | 20232.75 | 1 | 1 | 166 | 63.53333 |
| 48 | baseline_arm_1 | 6/23/2021 | 0.4 | 710 | 0 | 103 | 0 | 0 | 10 | 10 |
| 48 | 4_weeks_post_dose_arm_1 | 8/16/2021 | 0.4 | 950 | 390 | 302 | 0 | 0 | 10 | 10 |
| 48 | 12_weeks_post_dose_arm_1 | 10/14/2021 | 0.4 | 1560 | 180 | 270 | 0 | 0 | | |
| 48 | 24_weeks_post_dose_arm_1 | 1/20/2022 | 2.9 | 810 | 250 | 608 | 1 | 0 | | |
| 48 | bv_4_weeks_post_do_arm_2 | 2/24/2022 | 1.97 | 100 | 20 | 6856.25 | 1 | 0 | 10 | 10 |
| 48 | bv_12_weeks_post_d_arm_2 | 4/21/2022 | 0.4 | 5310 | 1980 | 19408 | 1 | 0 | | |

| | | | | | | | | | | |
|---|---|---|---|---|---|---|---|---|---|---|
| 48 | bv_24_weeks_post_d_arm_2 | 7/14/2022 | 31 | 20 | 260 | 7183.75 | 1 | 0 | 17 | 86.24667 |
| 48 | bv_48_weeks_post_d_arm_2 | 1/6/2023 | 26.64 | 810 | 890 | 10537.5 | 1 | 0 | | |
| 49 | baseline_arm_1 | 6/24/2021 | 0.4 | 30 | 40 | 213 | 1 | 0 | 14 | 10 |
| 49 | 4_weeks_post_dose_arm_1 | 8/17/2021 | 0.4 | 180 | 30 | 146 | 1 | 0 | 10 | 10 |
| 49 | 12_weeks_post_dose_arm_1 | 10/11/2021 | 0.4 | 70 | 20 | 145 | 1 | 0 | | |
| 49 | 24_weeks_post_dose_arm_1 | 1/12/2022 | 0.4 | No PHA | No PHA | 279 | 1 | 0 | | |
| 49 | bv_4_weeks_post_do_arm_2 | 2/28/2022 | 0.4 | 260 | 20 | 1764.58333 | 1 | 0 | 10 | 10 |
| 49 | bv_12_weeks_post_d_arm_2 | 5/6/2022 | 0.4 | 2700 | 1090 | 2004.33333 | 1 | 0 | | |
| 49 | bv_24_weeks_post_d_arm_2 | 7/18/2022 | 0.4 | 120 | 0 | 1687.08333 | 2 | 0 | 69.38 | 34.4 |
| 49 | bv_48_weeks_post_d_arm_2 | 1/9/2023 | 0.4 | 2130 | 1650 | 3113.75 | 2 | 0 | | |
| 31 | baseline_arm_1 | 4/8/2021 | 0.4 | 10 | 50 | 133 | 0 | 0 | | |
| 31 | 4_weeks_post_dose_arm_1 | 6/30/2021 | 0.4 | 2280 | 480 | 228 | 0 | 0 | 10 | 10 |
| 31 | 12_weeks_post_dose_arm_1 | 8/31/2021 | 0.4 | 50 | 0 | 68 | 0 | 0 | | |
| 31 | bv_48_weeks_post_d_arm_2 | 11/10/2021 | | 360 | 70 | | 0 | 0 | | |
| 31 | 24_weeks_post_dose_arm_1 | 11/23/2021 | 0.4 | 70 | 10 | 38 | 0 | 0 | | |
| 31 | bv_4_weeks_post_do_arm_2 | 1/3/2022 | 0.4 | 900 | 140 | 93 | 0 | 0 | | |
| 31 | bv_12_weeks_post_d_arm_2 | 3/4/2022 | 0.4 | 180 | 90 | 52 | 0 | 0 | 10 | 10 |
| 31 | bv_24_weeks_post_d_arm_2 | 6/2/2022 | 0.4 | 470 | 80 | 272 | 1 | 0 | 10 | 10 |
| 25 | baseline_arm_1 | 6/30/2021 | 0.4 | 0 | 0 | 107 | 0 | 0 | 10 | 14.42 |
| 25 | 4_weeks_post_dose_arm_1 | 11/23/2021 | 778.8 | 30 | 50 | 23705 | 0 | 0 | 400 | 10 |
| 25 | 12_weeks_post_dose_arm_1 | 2/24/2022 | 111 | 100 | 50 | 3274 | 1 | 0 | | |
| 25 | 24_weeks_post_dose_arm_1 | 4/22/2022 | 29.73 | 1520 | 2910 | 1820 | 1 | 0 | 10 | 10 |
| 25 | 48_weeks_post_dose_arm_1 | 9/28/2022 | | No PHA | No PHA | 7983.58333 | 1 | 0 | 10 | 829 |
| 3 | baseline_arm_1 | 6/30/2021 | 0.4 | 1610 | 150 | 1820 | 1 | 0 | 10 | 10 |
| 3 | 4_weeks_post_dose_arm_1 | 8/31/2021 | 206.9 | 1350 | 160 | 4536 | 1 | 0 | 53 | 10 |
| 50 | baseline_arm_1 | 2/22/2021 | 0.4 | 1397 | 53 | 605 | 1 | 0 | | |
| 50 | 4_weeks_post_dose_arm_1 | 7/1/2021 | 5628 | No PBMC | No PBMC | 29258 | 1 | 0 | 3663.5 | 47.52 |
| 50 | 12_weeks_post_dose_arm_1 | 8/27/2021 | 818.4 | 810 | 10 | 14881 | 1 | 0 | | |
| 50 | 24_weeks_post_dose_arm_1 | 11/22/2021 | 186.2 | 140 | 60 | 6576 | 1 | 0 | | |
| 50 | bv_4_weeks_post_do_arm_2 | 1/25/2022 | 25000 | 360 | 60 | 28123 | 1 | 0 | | |
| 50 | bv_12_weeks_post_d_arm_2 | 3/22/2022 | 25000 | 650 | 180 | 29725 | 1 | 0 | 9839 | 885.6 |
| 50 | bv_24_weeks_post_d_arm_2 | 6/13/2022 | 17782 | 250 | 60 | 26139 | 1 | 0 | 4164 | 973 |
| 50 | bv_48_weeks_post_d_arm_2 | 12/1/2022 | 7376 | 1030 | 690 | | 1 | 0 | | |
| 23 | baseline_arm_1 | 3/8/2021 | 0.4 | 0 | 0 | 79 | 0 | 0 | 10 | 10 |
| 23 | 4_weeks_post_dose_arm_1 | 7/1/2021 | 0.4 | 170 | 90 | 528 | 0 | 0 | 10 | 10 |
| 23 | 12_weeks_post_dose_arm_1 | 8/24/2021 | 0.4 | 30 | 130 | 430 | 0 | 0 | | |
| 23 | 24_weeks_post_dose_arm_1 | 12/1/2021 | 0.4 | 110 | 110 | 88.5 | 0 | 0 | | |
| 23 | bv_4_weeks_post_do_arm_2 | 3/18/2022 | 0.4 | 470 | 120 | 285.5 | 0 | 0 | 10 | 10 |
| 23 | bv_12_weeks_post_d_arm_2 | 5/3/2022 | 0.4 | 5820 | 2470 | 151.5 | 0 | 0 | 10 | 10 |

| | | | | | | | | | | |
|---|---|---|---|---|---|---|---|---|---|---|
| 45 | baseline_arm_1 | 4/8/2021 | 31.17 | 10 | 10 | 13264 | 1 | 0 | 461.55 | 39.98 |
| 45 | 4_weeks_post_dose_arm_1 | 7/2/2021 | 505.2 | 210 | 23 | 25114 | 1 | 0 | 2117 | 10 |
| 45 | 12_weeks_post_dose_arm_1 | 8/26/2021 | 469.2 | 3800 | 370 | 21775 | 1 | 0 | | |
| 45 | 24_weeks_post_dose_arm_1 | 11/18/2021 | 342.5 | 1320 | 140 | 12095 | 1 | 0 | | |
| 45 | bv_4_weeks_post_do_arm_2 | 1/7/2022 | 6643 | 5180 | 390 | 22733.8333 | 2 | 0 | 1093 | 30 |
| 45 | bv_12_weeks_post_d_arm_2 | 3/4/2022 | 6919 | 3300 | 630 | 26024.5833 | 2 | 0 | 1249 | 144.1 |
| 45 | bv_24_weeks_post_d_arm_2 | 5/20/2022 | 5282 | 2930 | 180 | 25603.0833 | 2 | 0 | 879.4667 | 140.9 |
| 45 | bv_48_weeks_post_d_arm_2 | 11/11/2022 | 6101 | 4930 | 160 | 24449.0833 | 2 | 0 | | |
| 60 | baseline_arm_1 | 4/19/2021 | 0.4 | 0 | 10 | 775 | 1 | 0 | 10 | 10 |
| 60 | 4_weeks_post_dose_arm_1 | 7/6/2021 | 2.57 | No response | No response | 25839 | 1 | 0 | 315.5 | 20 |
| 60 | 12_weeks_post_dose_arm_1 | 8/27/2021 | 0.5 | 0 | 0 | 20999 | 1 | 0 | | |
| 60 | 24_weeks_post_dose_arm_1 | 11/23/2021 | 2.51 | 80 | 110 | 18668 | 1 | 0 | | |
| 60 | bv_4_weeks_post_do_arm_2 | 5/25/2022 | 0.94 | 5100 | 2600 | 11981.25 | 1 | 0 | 10 | 10 |
| 60 | bv_12_weeks_post_d_arm_2 | 7/19/2022 | 0.64 | 630 | 220 | 7030.75 | 1 | 0 | | |
| 60 | bv_24_weeks_post_d_arm_2 | 10/17/2022 | | 130 | 190 | 3021.25 | 1 | 0 | 10 | 10 |
| 60 | bv_48_weeks_post_d_arm_2 | 4/10/2023 | 0.54 | 1460 | 480 | 2717 | 1 | 0 | | |
| 56 | baseline_arm_1 | 3/31/2021 | 0.4 | 57 | 10 | 178 | 1 | 0 | | |
| 56 | 4_weeks_post_dose_arm_1 | 7/12/2021 | 35.13 | 280 | 30 | 20207 | 1 | 0 | 35.255 | 10 |
| 56 | 12_weeks_post_dose_arm_1 | 9/8/2021 | 20.91 | 100 | 0 | 8618 | 1 | 0 | | |
| 56 | 24_weeks_post_dose_arm_1 | 12/2/2021 | 15.28 | 320 | 120 | 270 | 1 | 0 | | |
| 56 | bv_4_weeks_post_do_arm_2 | 3/3/2022 | 4690 | 300 | 100 | 17819 | 1 | 0 | | |
| 56 | bv_24_weeks_post_d_arm_2 | 7/22/2022 | 1929 | 170 | 0 | 13866 | 2 | 0 | 126 | 44.83 |
| 56 | bv_48_weeks_post_d_arm_2 | 2/1/2023 | 962.9 | 590 | 230 | | 2 | 0 | | |
| 17 | baseline_arm_1 | 3/5/2021 | 101.6 | 57 | 13 | 1883 | 1 | 0 | 73 | 10 |
| 17 | 4_weeks_post_dose_arm_1 | 8/23/2021 | 13077 | 60 | 0 | 14089 | 1 | 0 | 4358.5 | 192.5 |
| 17 | bv_4_weeks_post_do_arm_2 | 3/10/2022 | 8737 | 280 | 110 | 21247 | 1 | 0 | 2582 | 175.9333 |
| 17 | bv_12_weeks_post_d_arm_2 | 4/29/2022 | 4774 | 840 | 600 | 23670 | 1 | 0 | 1495 | 63 |
| 53 | baseline_arm_1 | 1/27/2021 | 0.4 | 97 | 0 | 116 | 0 | 0 | 10 | 10 |
| 53 | 4_weeks_post_dose_arm_1 | 10/5/2021 | 0.4 | 0 | 0 | 61 | 0 | 0 | 10 | 10 |
| 53 | 12_weeks_post_dose_arm_1 | 12/10/2021 | 0.4 | 40 | 20 | 119 | 0 | 0 | | |
| 53 | 24_weeks_post_dose_arm_1 | 3/8/2022 | 0.4 | 400 | 220 | 56 | 0 | 0 | | |
| 53 | bv_4_weeks_post_do_arm_2 | 5/2/2022 | 0.4 | 590 | 340 | 174.583333 | 1 | 0 | 10 | 10 |
| 53 | bv_12_weeks_post_d_arm_2 | 6/23/2022 | 0.4 | 530 | 300 | 1015.08333 | 1 | 0 | 10 | 10 |
| 53 | bv_24_weeks_post_d_arm_2 | 9/28/2022 | 4270 | No PHA | No PHA | 21259.5833 | 1 | 0 | 1227.65 | 185.8 |
| 53 | bv_48_weeks_post_d_arm_2 | 3/6/2023 | 2296 | | | | 1 | 0 | | |
| 26 | baseline_arm_1 | 9/9/2021 | 0.4 | 0 | 0 | 42 | 0 | 0 | 10 | 10 |
| 26 | 4_weeks_post_dose_arm_1 | 11/12/2021 | 0.4 | 10 | 0 | 728 | 0 | 0 | 10 | 10 |
| 26 | 12_weeks_post_dose_arm_1 | 1/21/2022 | 168.5 | 1580 | 220 | 17310 | 1 | 1 | | |
| 26 | 24_weeks_post_dose_arm_1 | 4/1/2022 | 6454 | 950 | 530 | 16835 | 1 | 1 | 282.9 | 193.8 |

| | | | | | | | | | | |
|---|---|---|---|---|---|---|---|---|---|---|
| 26 | 48_weeks_post_dose_arm_1 | 9/19/2022 | 1749 | 10 | 10 | | 1 | 1 | | |
| 11 | baseline_arm_1 | 9/23/2021 | 1.25 | No PBMC | No PBMC | 434.75 | 1 | 0 | 10 | 10 |
| 11 | 4_weeks_post_dose_arm_1 | 11/18/2021 | 268.2 | No PBMC | No PBMC | 8233 | 1 | 0 | 589 | 10 |
| 11 | 24_weeks_post_dose_arm_1 | 4/7/2022 | 84.28 | No PBMC | No PBMC | 3514 | 2 | 0 | | |
| 22 | baseline_arm_1 | 2/22/2021 | 0.4 | No response | No response | 58 | 0 | 0 | 10 | 28.49667 |
| 22 | 4_weeks_post_dose_arm_1 | 9/28/2021 | 327.9 | No PHA | No PHA | 759 | 0 | 0 | 240 | 10 |
| 22 | 12_weeks_post_dose_arm_1 | 11/22/2021 | 98.15 | No PBMC | No PBMC | 12108 | 0 | 0 | | |
| 22 | 24_weeks_post_dose_arm_1 | 2/22/2022 | 40.8 | 0 | 30 | 10839.8333 | 0 | 0 | | |
| 22 | 48_weeks_post_dose_arm_1 | 7/28/2022 | 27.72 | 180 | 20 | 8087.58333 | 1 | 0 | 10 | 10 |
| 21 | baseline_arm_1 | 3/31/2021 | 0.4 | 0 | 7 | 48 | 1 | 0 | 10 | 15.25 |
| 21 | 4_weeks_post_dose_arm_1 | 10/6/2021 | 36.9 | 90 | 30 | 4674 | 2 | 0 | 327 | 10 |
| 21 | 12_weeks_post_dose_arm_1 | 12/21/2021 | 8.57 | 150 | 20 | 2619 | 2 | 0 | | |
| 21 | 24_weeks_post_dose_arm_1 | 2/28/2022 | 3.83 | 390 | 80 | 8757.83333 | 2 | 0 | | |
| 21 | 48_weeks_post_dose_arm_1 | 8/17/2022 | 2.4 | 30 | 30 | 2455.58333 | 2 | 0 | 10 | 10 |
| 5 | baseline_arm_1 | 11/4/2021 | 0.4 | 10 | 0 | 152 | 0 | 0 | 10 | 10 |
| 5 | 4_weeks_post_dose_arm_1 | 12/29/2021 | 16.1 | 0 | 0 | 9464 | 0 | 0 | 30 | 10 |
| 5 | 12_weeks_post_dose_arm_1 | 2/23/2022 | 5.02 | 0 | 0 | 2530 | 0 | 0 | | |
| 16 | baseline_arm_1 | 5/4/2021 | 0.4 | 10 | 0 | 40 | 0 | 0 | 10 | 10 |
| 16 | 4_weeks_post_dose_arm_1 | 7/8/2021 | 0.4 | 380 | 110 | 42 | 0 | 0 | 16 | 10 |
| 16 | 12_weeks_post_dose_arm_1 | 9/8/2021 | 0.4 | 250 | 140 | 37 | 0 | 0 | | |
| 16 | bv_4_weeks_post_do_arm_2 | 10/14/2021 | 0.4 | 1520 | 100 | 37 | 0 | 0 | 10 | 10 |
| 16 | bv_12_weeks_post_d_arm_2 | 12/15/2021 | 0.4 | 700 | 90 | 340.833333 | 0 | 0 | | |
| 16 | bv_24_weeks_post_d_arm_2 | 3/14/2022 | 0.4 | 3530 | 700 | 280.833333 | 0 | 0 | 15 | 10 |
| 10 | baseline_arm_1 | 6/4/2021 | 0.4 | 40 | 20 | 82 | 0 | 0 | 10 | 10 |
| 10 | 4_weeks_post_dose_arm_1 | 7/26/2021 | 0.4 | 290 | 40 | 78 | 0 | 0 | 10 | 10 |
| 10 | 12_weeks_post_dose_arm_1 | 9/17/2021 | 0.4 | 310 | 40 | 68 | 0 | 0 | | |
| 10 | 24_weeks_post_dose_arm_1 | 12/7/2021 | 5945 | 2140 | 160 | 24293 | 1 | 1 | | |
| 13 | baseline_arm_1 | 6/11/2021 | 0.4 | 30 | 0 | 174.8 | 1 | 0 | 89.67 | 10 |
| 13 | 4_weeks_post_dose_arm_1 | 8/4/2021 | 0.4 | No PHA | No PHA | 569.6 | 1 | 0 | 10 | 10 |
| 13 | 12_weeks_post_dose_arm_1 | 9/28/2021 | 0.4 | 150 | 40 | 408.4 | 1 | 0 | | |
| 13 | 24_weeks_post_dose_arm_1 | 1/10/2022 | 1.21 | 700 | 270 | 157.9 | 1 | 0 | | |
| 51 | baseline_arm_1 | 6/14/2021 | 0.4 | 0 | 0 | 46 | 0 | 0 | 10 | 10 |
| 51 | 4_weeks_post_dose_arm_1 | 8/3/2021 | 0.4 | 120 | 0 | 367 | 0 | 0 | 10 | 10 |
| 51 | 12_weeks_post_dose_arm_1 | 10/13/2021 | 0.4 | 430 | 30 | 118 | 0 | 0 | | |
| 51 | 24_weeks_post_dose_arm_1 | 1/11/2022 | 0.4 | 170 | 30 | 30 | 0 | 0 | | |
| 51 | bv_4_weeks_post_do_arm_2 | 2/8/2022 | 0.4 | 1840 | 10 | 2971.58333 | 0 | 0 | 10 | 10 |
| 51 | bv_12_weeks_post_d_arm_2 | 4/20/2022 | 0.4 | 2610 | 460 | 2241.08333 | 0 | 0 | | |
| 51 | bv_24_weeks_post_d_arm_2 | 7/19/2022 | 0.4 | 150 | 30 | 3611.08333 | 1 | 0 | 15 | 24.51 |
| 51 | bv_48_weeks_post_d_arm_2 | 1/17/2023 | 0.4 | Low PHA, Hig | Low PHA, Hig | 5569 | 1 | 0 | | |

| | | | | | | | | | | |
|---|---|---|---|---|---|---|---|---|---|---|
| 2 | baseline_arm_1 | 6/17/2021 | 0.4 | 0 | 0 | 546 | 1 | 0 | 10 | 10 |
| 2 | 4_weeks_post_dose_arm_1 | 8/18/2021 | 0.4 | 850 | 30 | 38 | 1 | 0 | 10 | 10 |
| 14 | baseline_arm_1 | 6/22/2021 | 0.4 | 0 | 10 | 380 | 1 | 0 | 10 | 10 |
| 14 | 4_weeks_post_dose_arm_1 | 8/24/2021 | 0.45 | 200 | 30 | 452.9 | 1 | 0 | 10 | 10 |
| 14 | 12_weeks_post_dose_arm_1 | 10/28/2021 | 0.4 | No PHA | No PHA | 184.4 | 1 | 0 | | |
| 14 | 24_weeks_post_dose_arm_1 | 3/29/2022 | 0.4 | 3250 | 4320 | 377.6 | 2 | 0 | | |
| 37 | baseline_arm_1 | 6/22/2021 | 0.4 | 0 | 0 | 69 | 0 | 0 | 10 | 10 |
| 37 | 4_weeks_post_dose_arm_1 | 9/7/2021 | 139 | 30 | 0 | 20418 | 0 | 0 | 60 | 10 |
| 37 | 12_weeks_post_dose_arm_1 | 11/9/2021 | 158 | 30 | Plating error | 10995.5833 | 0 | 0 | | |
| 37 | 24_weeks_post_dose_arm_1 | 2/8/2022 | 73.86 | 10 | 30 | 5337.4 | 0 | 0 | | |
| 37 | 48_weeks_post_dose_arm_1 | 9/27/2022 | 2131 | 80 | 10 | 10597.3333 | 1 | 0 | 311.7 | 32.1 |
| 4 | baseline_arm_1 | 7/6/2021 | 0.4 | 0 | 0 | 56 | 0 | 0 | 10 | 10 |
| 4 | 4_weeks_post_dose_arm_1 | 8/25/2021 | 0.4 | 140 | 20 | 559 | 0 | 0 | 31.65 | 10 |
| 4 | 12_weeks_post_dose_arm_1 | 10/19/2021 | 0.4 | 300 | 100 | 4562 | 0 | 0 | | |
| 35 | baseline_arm_1 | 7/13/2021 | 0.4 | 20 | 0 | 348 | 1 | 0 | 10 | 10 |
| 35 | 4_weeks_post_dose_arm_1 | 9/13/2021 | 1.76 | 410 | 50 | 5288 | 1 | 0 | 71 | 10 |
| 35 | 12_weeks_post_dose_arm_1 | 11/16/2021 | 1.04 | 220 | 40 | 2012 | 1 | 0 | | |
| 35 | 24_weeks_post_dose_arm_1 | 2/8/2022 | 0.54 | 270 | 10 | 1345.6 | 2 | 0 | | |
| 35 | 48_weeks_post_dose_arm_1 | 9/19/2022 | 0.4 | 40 | 10 | 502 | 2 | 0 | 10 | 10 |
| 42 | baseline_arm_1 | 8/3/2021 | 0.4 | 0 | 0 | 46 | 0 | 0 | 10 | 10 |
| 42 | 4_weeks_post_dose_arm_1 | 10/11/2021 | 136 | 90 | 10 | 74 | 0 | 0 | 161 | 10 |
| 42 | 12_weeks_post_dose_arm_1 | 11/17/2021 | 83.11 | 130 | 30 | 12773 | 0 | 0 | | |
| 42 | bv_4_weeks_post_do_arm_2 | 3/14/2022 | 1942 | 1040 | 330 | 13087.8333 | 0 | 0 | 248 | 18 |
| 42 | bv_12_weeks_post_d_arm_2 | 5/11/2022 | 731.9 | 400 | 110 | 9428.83333 | 0 | 0 | | |
| 42 | bv_24_weeks_post_d_arm_2 | 7/19/2022 | 130.5 | 220 | 10 | 5750.33333 | 0 | 0 | 15 | 10 |
| 42 | bv_48_weeks_post_d_arm_2 | 1/24/2023 | 24.8 | 170 | 40 | 10453 | 0 | 0 | | |
| 32 | baseline_arm_1 | 8/25/2021 | 0.4 | 10 | 0 | 108 | 0 | 0 | 10 | 10 |
| 32 | 4_weeks_post_dose_arm_1 | 10/19/2021 | 0.4 | 20 | 80 | 116 | 0 | 0 | 10 | 10.69 |
| 32 | 12_weeks_post_dose_arm_1 | 12/17/2021 | 0.4 | 130 | 40 | 117 | 0 | 0 | | |
| 32 | 24_weeks_post_dose_arm_1 | 3/18/2022 | 0.4 | 0 | 0 | 148 | 0 | 0 | | |
| 32 | bv_4_weeks_post_do_arm_2 | 5/10/2022 | 0.4 | 180 | 40 | 39 | 0 | 0 | 10 | 10 |
| 32 | bv_12_weeks_post_d_arm_2 | 7/8/2022 | 0.4 | 340 | 350 | 36 | 0 | 0 | | |
| 32 | bv_24_weeks_post_d_arm_2 | 9/27/2022 | 0.4 | 250 | 70 | 30 | 1 | 0 | 10 | 10 |
| 6 | baseline_arm_1 | 5/6/2021 | 0.4 | 0 | 0 | 61 | 0 | 0 | 22.4 | 10 |
| 6 | 4_weeks_post_dose_arm_1 | 6/22/2021 | | 190 | 20 | 64 | 0 | 0 | 50 | 10 |
| 6 | 12_weeks_post_dose_arm_1 | 8/24/2021 | 0.4 | 120 | 10 | 53 | 0 | 0 | | |
| 39 | baseline_arm_1 | 9/7/2021 | 0.4 | 20 | 0 | 77 | 0 | 0 | 10 | 10 |
| 39 | 4_weeks_post_dose_arm_1 | 10/26/2021 | 1.33 | 610 | 110 | 4148.4 | 0 | 0 | 10 | 10 |
| 39 | 12_weeks_post_dose_arm_1 | 12/28/2021 | 6.45 | 550 | 10 | 896 | 0 | 0 | | |

| | | | | | | | | | | |
|---|---|---|---|---|---|---|---|---|---|---|
| 39 | bv_4_weeks_post_do_arm_2 | 2/7/2022 | 1827 | 2820 | 10 | 150 | 0 | 0 | 314.2333 | 23.87 |
| 39 | bv_12_weeks_post_d_arm_2 | 3/24/2022 | 884 | 1870 | 1770 | 12842 | 0 | 0 | | |
| 39 | bv_24_weeks_post_d_arm_2 | 7/5/2022 | 550.4 | 2150 | 190 | 14102 | 1 | 0 | 85.06 | 46.75 |
| 39 | bv_48_weeks_post_d_arm_2 | 1/17/2023 | 136 | Low PHA, Hig | Low PHA, Hig | 12637 | 1 | 0 | | |
| 33 | baseline_arm_1 | 9/8/2021 | 0.4 | 20 | 10 | 97 | 0 | 0 | 10 | 10 |
| 33 | 4_weeks_post_dose_arm_1 | 11/8/2021 | 0.4 | 50 | 20 | 962 | 0 | 0 | 10 | 10 |
| 33 | 12_weeks_post_dose_arm_1 | 1/12/2022 | 119.7 | 410 | 30 | 1926 | 1 | 1 | | |
| 33 | 24_weeks_post_dose_arm_1 | 5/3/2022 | 1029 | 150 | 120 | 26083 | 1 | 1 | | |
| 33 | 48_weeks_post_dose_arm_1 | 11/2/2022 | | 180 | 170 | 806 | 1 | 1 | 30.61 | 10 |
| 34 | baseline_arm_1 | 9/15/2021 | 0.4 | 10 | 0 | 150 | 0 | 0 | 63.31 | 10 |
| 34 | 24_weeks_post_dose_arm_1 | 6/24/2022 | 5086 | 190 | 50 | 11173.4167 | 1 | 1 | | |
| 34 | 48_weeks_post_dose_arm_1 | 12/20/2022 | | Low PHA, Hig | Low PHA, Hig | 12848 | 1 | 1 | | |
| 12 | baseline_arm_1 | 5/7/2021 | 1.33 | 40 | 0 | 410 | 1 | 0 | 10 | 11 |
| 12 | 4_weeks_post_dose_arm_1 | 7/13/2021 | 31.84 | 510 | 30 | 1000 | 1 | 0 | 66 | 10 |
| 12 | 12_weeks_post_dose_arm_1 | 9/15/2021 | 33.2 | 110 | 30 | 1123 | 1 | 0 | | |
| 12 | 24_weeks_post_dose_arm_1 | 12/8/2021 | 31.36 | 2490 | 200 | 1114 | 1 | 0 | | |
| 38 | baseline_arm_1 | 5/7/2021 | 0.4 | 0 | 0 | 48 | 0 | 0 | | |
| 38 | 4_weeks_post_dose_arm_1 | 7/13/2021 | 0.4 | 10 | 0 | 46 | 0 | 0 | 10 | 10 |
| 38 | 12_weeks_post_dose_arm_1 | 8/31/2021 | 0.4 | 0 | 0 | 62 | 0 | 0 | | |
| 38 | bv_24_weeks_post_d_arm_2 | 3/1/2022 | 0.4 | 0 | 0 | 60 | 0 | 0 | 20 | 10 |
| 38 | bv_48_weeks_post_d_arm_2 | 9/6/2022 | 0.4 | No PHA | No PHA | 3043 | 0 | 0 | 10 | 10 |
| 8 | baseline_arm_1 | 5/11/2021 | 0.4 | 0 | 0 | 39 | 0 | 0 | 23.91 | 10 |
| 8 | 4_weeks_post_dose_arm_1 | 6/28/2021 | 0.4 | 30 | 0 | 88 | 0 | 0 | 10 | 10 |
| 8 | 12_weeks_post_dose_arm_1 | 8/25/2021 | 0.4 | 50 | 0 | 44 | 0 | 0 | | |
| 8 | bv_4_weeks_post_do_arm_2 | 10/27/2021 | 0.4 | 300 | 10 | 48 | 0 | 0 | 10 | 10 |
| 9 | baseline_arm_1 | 5/11/2021 | 0.4 | 0 | 0 | 75 | 0 | 0 | 10 | 10 |
| 9 | 4_weeks_post_dose_arm_1 | 8/4/2021 | 0.4 | 60 | 0 | 11 | 0 | 0 | 10 | 10 |
| 9 | 24_weeks_post_dose_arm_1 | 11/30/2021 | 0.4 | 870 | 350 | 3657 | 1 | 0 | | |
| 1 | baseline_arm_1 | 5/18/2021 | 0.4 | No PBMC | No PBMC | 208 | 1 | 0 | 10 | 10 |
| 1 | 4_weeks_post_dose_arm_1 | 8/18/2021 | 241 | 20 | 0 | 11477 | 1 | 0 | 329.1 | 10 |
| 15 | baseline_arm_1 | 6/4/2021 | 0.4 | 0 | 0 | 90 | 0 | 0 | 10 | 10 |
| 15 | 4_weeks_post_dose_arm_1 | 7/28/2021 | 0.4 | No PHA | No PHA | 144 | 0 | 0 | 14 | 10 |
| 15 | bv_4_weeks_post_do_arm_2 | 10/6/2021 | 5061 | 30 | 70 | 8423 | 0 | 0 | | |
| 15 | bv_24_weeks_post_d_arm_2 | 3/8/2022 | 587.2 | 130 | 700 | 2113 | 0 | 0 | 499.6 | 124.4 |
